# Continuous Indexing of Fibrosis (CIF): Improving the Assessment and Classification of MPN Patients

**DOI:** 10.1101/2022.06.06.22276014

**Authors:** Hosuk Ryou, Korsuk Sirinukunwattana, Alan Aberdeen, Gillian Grindstaff, Bernadette Stolz, Helen Byrne, Heather A. Harrington, Nikolaos Sousos, Anna L. Godfrey, Claire N. Harrison, Bethan Psaila, Adam J. Mead, Gabrielle Rees, Gareth D.H. Turner, Jens Rittscher, Daniel Royston

**Author notes:** Corresponding author: Daniel Royston (, Nuffield Division of Clinical Laboratory Sciences, University of Oxford, Oxford).

## Abstract

The detection and grading of fibrosis in myeloproliferative neoplasms (MPN) is an important component of disease classification, prognostication and disease monitoring. However, current fibrosis grading systems are only semi-quantitative and fail to capture sample heterogeneity. To improve the detection, quantitation and representation of reticulin fibrosis, we developed a machine learning (ML) approach using bone marrow trephine (BMT) samples (n = 107) from patients diagnosed with MPN or a reactive / nonneoplastic marrow. The resulting Continuous Indexing of Fibrosis (CIF) enhances the detection and monitoring of fibrosis within BMTs, and aids the discrimination of MPN subtypes. When combined with megakaryocyte feature analysis, CIF discriminates between the frequently challenging differential diagnosis of essential thrombocythemia (ET) and pre-fibrotic myelofibrosis (pre-PMF) with high predictive accuracy [area under the curve = 0.94]. CIF also shows significant promise in the identification of MPN patients at risk of disease progression; analysis of samples from 35 patients diagnosed with ET and enrolled in the Primary Thrombocythemia-1 (PT-1) trial identified features predictive of post-ET myelofibrosis (area under the curve = 0.77). In addition to these clinical applications, automated analysis of fibrosis has clear potential to further refine disease classification boundaries and inform future studies of the micro-environmental factors driving disease initiation and progression in MPN and other stem cell disorders. The image analysis methods used to generate CIF can be readily integrated with those of other key morphological features in MPNs, including megakaryocyte morphology, that lie beyond the scope of conventional histological assessment.

**Key Points:**

- Machine learning enables an objective and quantitative description of reticulin fibrosis within the bone marrow of patients with myeloproliferative neoplasms (MPN),
- Automated analysis and Continuous Indexing of Fibrosis (CIF) captures heterogeneity within MPN samples and has utility in refined classification and disease monitoring
- Quantitative fibrosis assessment combined with topological data analysis may help to predict patients at increased risk of progression to post-ET myelofibrosis, and assist in the discrimination of ET and pre-fibrotic PMF (pre-PMF)

## Introduction

Reticulin fibers, comprising type III collagen, are an important component of the normal bone marrow extracellular matrix (ECM) essential for the maintenance of hematopoiesis. In normal marrow, silver impregnation techniques highlight the reticulin substrate as a delicate network of thin, uniform fibers coursing through the intertrabecular spaces, with variable condensation around small blood vessels. In several pathological conditions, including acute lymphoblastic leukemia and myelodysplastic syndrome, this organized reticulin meshwork is perturbed, with increasing fiber quantity, thickness and intersections accompanied by the deposition of additional collagen subtypes with disease progression^1-3^. However, the diagnostic and prognostic importance of reticulin meshwork is best characterized in the Philadelphia-negative myeloproliferative neoplasms (MPNs), a group of disorders in which acquired mutations in hematopoietic stem cells affect the MPL-JAK-STAT signaling pathway and drive excessive proliferation of ≥1 blood lineage^4-7^. Although the precise mechanisms driving marrow fibrosis remain poorly understood, it is increasingly recognized that the initiation and progression of fibrosis in MPNs reflects a pathological cytokine / chemokine-driven inflammatory response to clonal myeloproliferation, driven by neoplastic hematopoietic stem cells (HSC)^8-10^.

The importance of fibrosis estimation in MPNs is embedded in the current World Health Organization (WHO) classification scheme of the common MPNs: essential thrombocythemia (ET), polycythemia vera (PV), primary myelofibrosis (PMF) and pre-fibrotic primary myelofibrosis (pre-PMF)^11^. Fibrosis severity also has clinical implications in MPNs, with minor fibrosis in PV associated with inferior survival and more advanced fibrosis associated with a complex karyotype^12-14^. In PMF, increasing fibrosis is associated with worsening hematological, clinical and molecular parameters and overall prognosis^15-17^. The latest version of the WHO fibrosis scoring system comprises four categories (MF 0-3). Grade MF-0 is defined as scattered linear reticulin, with no intersections (typical of normal marrow); MF-1 as a loose network of reticulin, with many intersections; MF-2 as a diffuse and dense increase in reticulin, with extensive intersections and occasional focal bundles of collagen and / or focal osteosclerosis; and, MF-3 as a diffuse and dense increase in reticulin, with extensive intersections and coarse bundles of collagen, often associated with significant osteosclerosis. Although these grade descriptions are qualitative and subjective, several studies have demonstrated reasonable-to-good concordance of grade assignment between specialist hematopathologists^18-21^. Nonetheless, the WHO grading scheme fails to accommodate fibrotic heterogeneity within BMT specimens and cannot capture the spectrum of fibrosis within existing grade categories.

In response, we sought to develop an automated machine learning (ML) methodology to objectively quantify reticulin fibrosis using routinely prepared reticulin-stained BMT samples. This new quantitative approach was designed to overcome the limitations of the current WHO grading system that fails to capture subtle, focal fibrotic foci and the spectrum of fibrosis within grade categories. Manually annotated regions of fibrosis corresponding to each WHO grade were used to create a set of image tiles from which an initial model of fibrosis severity was trained. This model was iteratively refined on a set of locally sourced normal / reactive (n = 12) and MPN samples (n = 95) spanning the range of fibrosis encountered in clinical practice and incorporating each MPN subtype. Specialist hematopathologist review assisted the creation of a ranked-list of fibrosis severity in which each image tile receives a predicted fibrosis score between 0 and 1 (Continuous Index of Fibrosis [CIF]). The predicted scores of new, unseen tiles were then converted to a quantitative fibrosis map overlaid onto whole sample images. Analysis of MPN sample cohorts allowed us to capture the spectrum and heterogeneity of fibrosis within established MPN and normal / reactive BMT samples. We hypothesized that such an approach would enhance the accuracy of fibrosis assessment in MPN samples, with implications for improved disease classification (particularly distinction of ET and pre-PMF) and refined descriptions of disease monitoring / response to therapy. To assess the potential for improving disease prognostication, we also applied our methodology to a set of well characterized ET patients (n = 37) enrolled to a large multi-centre clinical trial (PT-1) with long term clinical follow and evidence of either stable disease or progression to post-ET myelofibrosis.

## Materials and Methods

An overview of the methodologies employed in this study is given as Figure 1.

**Figure 1.**
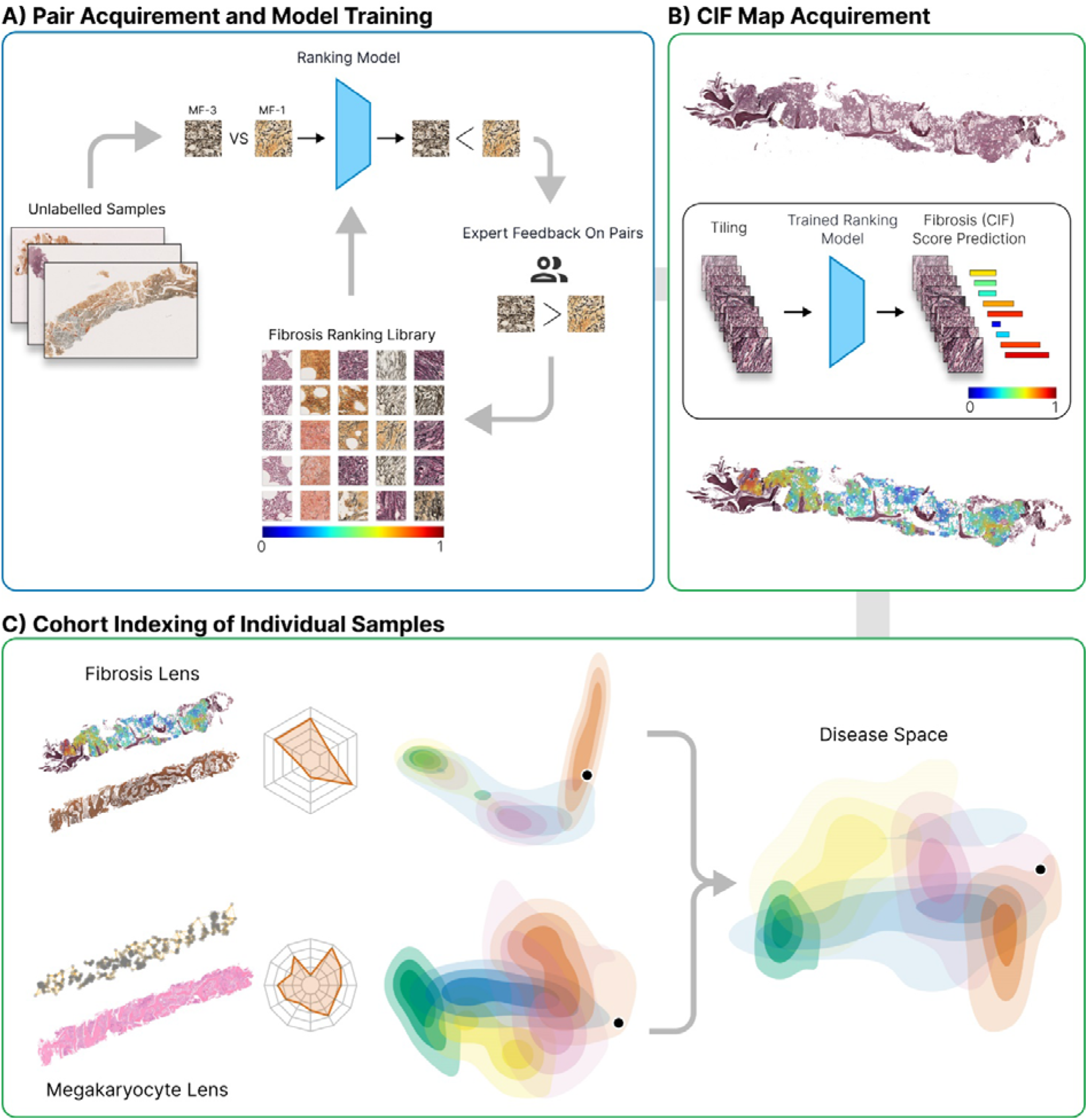
Overview of the computational steps for detection, quantitation and visualization of reticulin fibrosis in BMTs. (A) Image tiles are extracted from manually annotated (segmented) areas of fibrosis and labeled with the corresponding grade. Extracted tiles are then used to train a ranking-CNN (convolutional neural network) model to output scores using a pairwise sample approach, with higher scores corresponding to more severe fibrosis. Based on the initial trained model, we adopted a human-in-the-loop approach for manual image ranking. (B) With the finalized model, a CIF map of each sample is acquired by predicting the scores of all extracted tiles and visualizing this as a colormap superimposed upon the original reticulin-stained image. (C) From the predicted CIF scores and measurements of tile distribution, the fibrosis features are represented in 2-dimensional disease space to allow comparison of MPN subtypes and indexing of individual patient BMTs against a sample library. Fibrosis features can then be combined with those of other BMT constituents (e.g. megakaryocytes) to refine the disease space.

### Clinical samples

BMT samples were obtained from the archive of OUH NHS Foundation Trust. All specimens were of sufficient technical quality for use in conventional histological reporting. Samples were fixed in 10% neutral buffered formalin for 24 hours prior to decalcification in 10% EDTA for 48 hours. Whole slide scanned images (Hamamatsu NanoZoomer 2.0HT / 40X / NDPI files or 3DHISTECH 250 Flash III Dx / 40X / MRXS files) were prepared from 2-3 μm reticulin-stained (Gomori and Sweet) sections cut from formalin-fixed paraffin-embedded (FFPE) blocks. The data set comprised 107 samples (36 ET, 19 PV, 23 MF, 17 pre-PMF, and 12 reactive / nonneoplastic) with “reactive” samples sourced from patients in whom there was no evidence of bone marrow malignancy, persistent thrombocytosis or underlying myeloid disorder. ET, PV and MF (primary [PMF] or secondary [SMF]) samples were obtained from patients in whom this was either an established or new diagnosis, satisfying the diagnostic criteria of the current WHO classification (2016), and were designated following review by a myeloid multidisciplinary meeting (MDM). BMTs were identified from the laboratory reporting system or MDM records. A summary of the key patient characteristics is provided in supplemental Table 1. Additional MPN samples (n = 35) were obtained from the Primary Thrombocythemia-1 (PT-1) trial cohort; a multicentre international trial in ET in which newly diagnosed and previously treated patients with ET, aged 18 years or over, were recruited into one of three studies (previously published) depending on their risk of vascular complications^22-24^. The driver mutation status for all MPN samples is summarized in supplemental Figure 1. This work was conducted as part of the INForMeD study (INvestigating the genetic and cellular basis of sporadic and Familial Myeloid Disorders; IRAS ID: 199833; REC reference: 16/LO/1376; PI: Prof AJ Mead) with all patients having provided written informed consent.

### Automated identification of fibrosis and severity assessment

Reticulin staining employs a silver impregnation technique that highlights reticulin fibers as black linear extracellular material, although minor variations in routine laboratory practice (including counterstaining and toning) impart a range of color to bone and cellular elements. Therefore, digitized reticulin images were converted into grayscale to prevent any color variation adversely influencing our model’s performance. Grayscale transformation is employed in texture analysis^25,26^, and is well suited to silver-stained images in which the monochrome properties of the black reticulin fibers are largely unaffected by variations in background color. Two sets of BMT samples capturing the spectrum of marrow fibrosis in MPN (MF-0 to MF-3) were used for the training (39 samples) and validation (18 samples) steps of our model generation. For the initial training and validation stages, uniformly sized tiles (512 × 512 pixels [0.22 □m per pixel]) were extracted from manually annotated samples prepared by a specialist hematopathologist (DR) and deemed suitable for fibrosis estimation. Sample areas that contained significant amounts of bone, crush artifacts or hemorrhage were excluded. As bone is a significant component of BMT samples, a deep learning model based on UNet^27^ was used to assist in the segmentation and exclusion of bony trabeculae (for implementation and hyperparameter settings see supplemental Table 2). For subsequent rounds of training and validation employing a human-in-the-loop approach, a sliding window of 512 × 512 pixels with a stride of 256 pixels (supplemental Figure 2), was used to extract tiles that satisfied each of three criteria: fat regions account for <50% of tile area; bone or bone fragments account for <1% of tile area; and, blood vessels account for <10% of tile area. We reasoned that this rule set for tile extraction and analysis was an acceptable compromise, maximizing the analyzable area of each sample while adhering to the convention of restricting fibrosis grading to areas of hematopoiesis (excluding peritrabecular and perivascular tissues).

Significant barriers to developing an automated approach to fibrosis grading include the absence of objective criteria for assessing fibrosis severity, and a means for combining the distinct features of reticulin quantity, fiber thickness and intersections currently employed in the WHO fibrosis grading system. We also recognized the importance of developing an automated method that accommodates a continuous spectrum of fibrosis severity within and between MF grades, and that a simple classification model and regression approach to assign fibrosis scores is inadequate. Therefore, we employed a Learning to Rank (LTR) strategy called RankNet to train a model that estimates sample fibrosis in the form of an ordered ranking of feature severity^28^. RankNet is an established LTR algorithm ideally suited to tasks in which the ordering of items based upon a set of features is more important than an exact score or classification. This RankNet approach was then combined with a Convolutional Neural Network (CNN) to build a Ranking-CNN model^29^. A CNN was used as they demonstrate good performance in medical image analysis tasks^27,30^ (for implementation and hyperparameter settings see supplemental Table 3). To determine the ground truth of analyzed images, a pairwise ranking strategy suitable for rapid and intuitive human review was adopted, with three specialist hematopathologists selecting the most severe of two candidate image regions using conventional WHO fibrosis criteria. For image pairs resulting in discordant ranking between pathologists, a majority rule was followed. (Please refer to supplemental Figure 3 for an overview of the initial tile pair acquisition and model training). Since the output scores of the ranking-CNN model are continuous and with no limit, and our objective was to estimate the severity of fibrosis, we used the range of the predicted scores based on the training set to normalize the output scores between 0 and 1. This range of the normalized score was used as the reference of the severity of the fibrosis, with scores (Continuous Index of Fibrosis [CIF] scores) approaching 1 representing the most severe fibrosis.

We adopted a human-in-the-loop approach for manual image ranking (supplemental Figure 4) as this minimized the number of pairwise image comparison tasks required for each iteration of model training and validation. Briefly, in each round the trained model was used to predict the ranking for unseen tiles. A subset of pairs were reviewed and a rank label was determined by three hematologists (see supplemental Methods: Training of the ranking-CNN model). We created 8307 pairs for the training set and 3063 pairs for the validation set across three rounds of manual ranking (supplemental Table 4). Please refer to supplemental Figure 5 for examples of image pairs equating to each fibrosis grade.

### Generating image maps of fibrosis severity and feature extraction

Generating fibrosis severity maps based on our CIF model output score is an efficient and intuitive method of visualizing fibrosis throughout a sample. To acquire these CIF maps, a sliding window of 512 × 512 pixels was used within the annotated region, with a stride of 256 pixels. The model outputs a score for each window, with regions of overlap receiving an average score of the overlapping windows.

To allow comparison between MPN subtypes and normal / reactive marrow, three sets of features relating to the analyzed tiles extracted from each sample were used: average CIF score, tile distribution, and Shannon entropy (or Shannon diversity index) of tile distribution. The average score is correlated to the overall sample fibrosis, with Shannon entropy and tile distribution related to the population diversity of the extracted tile scores. Shannon entropy (henceforth referred to as heterogeneity) captures the ‘unevenness’ of tile scores; increasing values correspond to a uniform distribution of tile scores across the entire range of possible scores. The tile distribution reflects the extent to which particular tile CIF scores are enriched in each sample. As the output CIF scores from our model were continuous between 0 and 1, scores were divided into four groups or ‘bins’ that broadly corresponded to the four WHO fibrosis grades of MF-0, MF-1, MF-2 and MF-3. Using images extracted from the previously manually annotated regions (training set), the data was first balanced by having equal numbers of tiles drawn from areas manually assigned to each of the four MF grades. Our model was then applied to this balanced dataset, with the predicted scores normalized and ranked from the lowest to highest score. The range of the normalized scores for each of the four bins was then determined by dividing the total number of tiles into four equally sized sets corresponding to each bin. The difference in fibrosis between MPN subtypes was calculated via the Mann-Whitney-Wilcoxon test where p-value (*P*) < 0.05 is considered statistically significant.

### Topological data analysis of ET and pre-PMF samples

To interrogate the relationship between fibrotic foci within ET and pre-PMF samples, we employed topological data analysis (TDA), a relatively new field in computational mathematics that studies the shape and connectivity of data^31,32^. TDA has previously found application in quantifying the spatial patterns of various cell distributions within cancer tissue^33^. Persistent homology, a prominent and robust TDA algorithm^34^, enabled us to explore the connectivity pattern of fibrotic foci across a continuous range of spatial scales within our samples. The input for this analysis was the CIF tile scores and the output was a multiscale summary of the spatial connectivity of the CIF scores called a barcode, a topological fingerprint generated using Python Ripser version 0.6.2^35^ (Figure 5). The barcode tracks the persistence and connectivity of fibrotic foci as they appear and evolve in the image at different threshold values of the CIF score. Quantitative properties of the barcode could then be used for subsequent analysis and classification. To distinguish between ET and pre-PMF samples, a random forest classifier was then applied in Python, using the package scikit-learn^36^, with a classifier comprising 100 decision trees. The importance of individual features was assessed using Gini importance^37,38^. For further details and a description of the topological statistics used for this analysis, please refer to supplemental Methods: Topology data analysis.

## Results

### Estimation of BMT fibrosis severity using a ranked list approach

We employed a human-in-the-loop methodology to efficiently build a ranked list of fibrosis severity comprising 11448 image tiles extracted from reticulin-stained BMT sections. Following an initial round of pairwise ranking using tiles extracted from manually annotated whole slide-images, two subsequent rounds of manual ranking were fed back into the ML model for further training. The average manual ranking concordance by three hematopathologists after the first round of training and validation was high (88.40%; supplemental Table 5). Unsurprisingly, concordance was highest between extremes of MF grade (e.g. MF-0 vs MF-3), with significant discordance largely restricted to pair samples extracted from areas manually segmented as being within the same WHO fibrosis grade (e.g. MF-2 vs MF-2). After three rounds of training and validation, our fibrosis ranking model achieved 93.99% accuracy (see supplemental Tables 6 and 7 for the ranking performance within different image pairs and interobserver agreement).

As expected, the predicted tile ranking output of our model ranked highly fibrotic sample areas as those containing numerous thick reticulin fibers and bundles with frequent intersections (Figure 2B; supplemental Figure 6). Our model appeared to consistently rank tiles specifically on the reticulin fiber properties, with detailed visual inspection revealing no evidence of any significant impact resulting from non-specific or artifactual staining properties.

**Figure 2.**
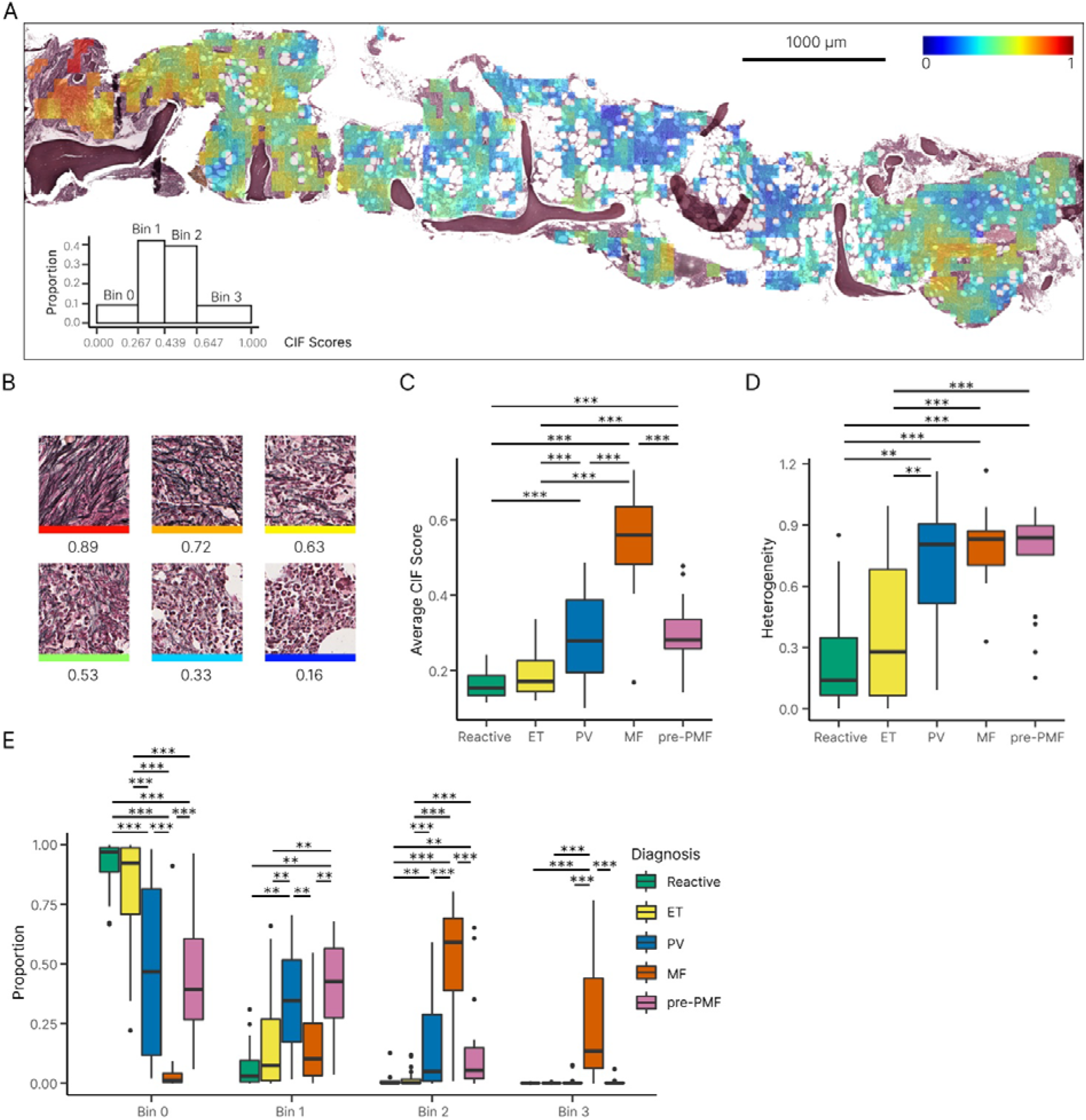
Distribution of tile CIF scores across MPN and reactive samples. (A) Example of a false-colored fibrosis heatmap, with CIF scores overlaid onto the original reticulin-stained BMT image [summary of tile distribution in bottom left]. (B) Individual tiles receive a CIF score from 0 to 1 depending on the severity of reticulin fibrosis, with high scores assigned to tiles displaying increased fiber quantity, thickness and intersections. Boxplots of the (C) average CIF score and (D) heterogeneity of tile distribution for MPN and reactive samples. (E) Boxplot for the distribution of CIF scores (grouped into bins of increasing fibrosis) for MPN and reactive / normal samples.

### Visualization of fibrosis using CIF image mapping

To better understand the output of the ranking model, we sought to convert the derived CIF tile scores into an intuitive representation suitable for assessment by hematopathologists. The normalized CIF scores were converted to a color scale that could be superimposed upon a BMT image to generate a false-colored image (Figure 2A; Figure 3). This representation allows an intuitive interpretation of the model output and contextualizes the results against familiar morphological tissue landmarks.

**Figure 3.**
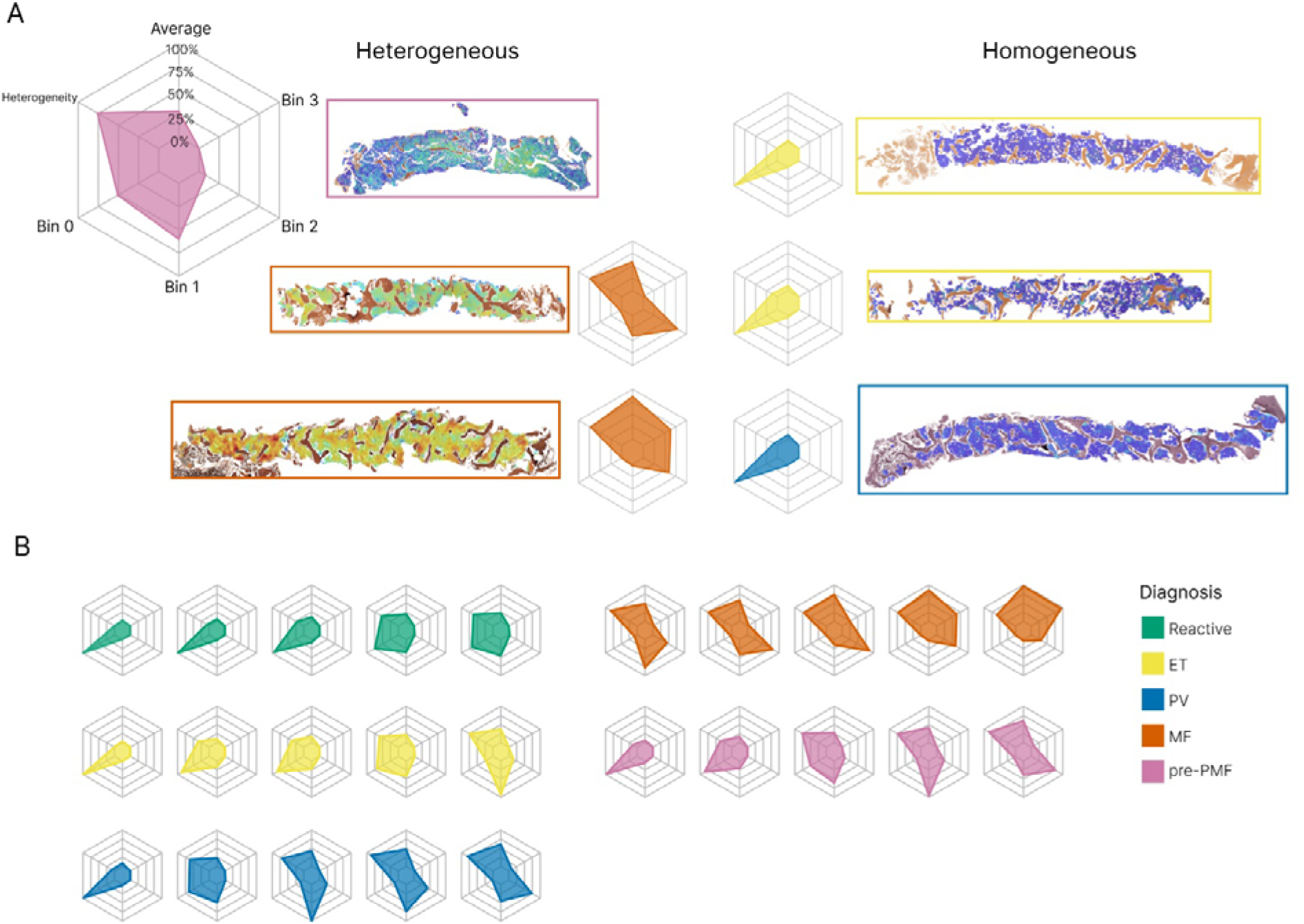
Variability of reticulin fibrosis within BMT samples. (A) Radar plots capture the average CIF tile score, tile distribution across the four bins (or ranges), and heterogeneity of tile distribution. Examples of homogenous and heterogenous patterns of fibrosis are shown. (B) Examples of radar plots for reactive samples and each MPN subgroup.

### Heterogeneity of reticulin fibrosis in BMT samples is associated with MPN subtype

To accurately and objectively compare fibrosis quantitation between MPN subtypes and reactive samples, we determined average whole sample statistics that captured fibrosis severity and heterogeneity (Figure 2C-D; Figure 3). As expected, MF samples demonstrated significantly more fibrosis (average CIF score) than other MPN subtypes or the reactive / normal marrows. Equally expected was the finding of no significant difference between the average fibrosis scores in ET and reactive / normal samples; areas of minimal fibrosis amounting to MF-1 being well described in healthy marrow. In keeping with previous descriptions of patchy and variable fibrosis in PV, the average fibrosis score for PV was moderately higher than that of ET (PV 0.30 vs ET 0.19, *P =* 5E-4). Average fibrosis scores for PV and pre-PMF were identical (PV 0.30 vs pre-PMF 0.30, *P* = 0.94). Of particular interest, pre-PMF samples contained a significantly higher average CIF score than ET (pre-PMF 0.30 vs ET 0.19, *P* = 8E-5), despite meeting the diagnostic requirement of containing ≤ WHO grade MF-1 by conventional histological assessment. Given the importance of BMT histological assessment in distinguishing patients with ET and pre-PMF, this result suggested that our automated fibrosis analysis may have clinical utility in resolving this frequently challenging differential diagnosis.

In order to determine the distribution of the tile scores for each MPN subtype, we subdivided the tile CIF scores into four distinct groups, or bins, that broadly correspond to each of the four established WHO fibrosis grade categories from the initial segmentation set (Figure 2E). As expected, MF cases accounted for almost all of the tiles assigned to bin 3, although less fibrotic / non-fibrotic tiles were also frequently encountered in MF samples. Also expected was the observation that ET samples predominantly comprised tiles from bins 0 and 1 (82.36% and 16.10%, respectively), broadly similar to the reactive / normal samples, consistent with fibrosis in ET seldom exceeding focal areas of conventional WHO grade MF-1 (supplemental Table 8). The PV samples contained a fairly wide distribution of tiles from bins 0-2, with significantly more tile scores allocated to bin 1 than ET (PV 0.35 vs ET 0.16, *P =* 0.002) and bin 2 (PV 0.17 vs ET 0.02, *P* = 5E-4). Notably, samples of pre-PMF contained tile scores that were significantly different from those of ET; despite being predominantly composed of tiles assigned to bins 0 and 1 (46.52% and 38.85%, respectively), tile scores assigned to bin 2 were significantly increased in the pre-PMF samples (pre-PMF 14.16% vs ET 1.51%, *P* = 2E-4), although areas of obvious WHO grade MF-2 (as determined by routine histological review) were absent from these samples in line with current WHO diagnostic criteria (supplemental Table 8). Tile score distributions observed for the PV and pre-PMF samples were not significantly different. Of note, fibrosis heterogeneity did not appear to be simply correlated to average CIF scores, with no significant difference observed between the fibrosis heterogeneity of MF, PV and pre-PMF samples (MF 0.80 vs PV 0.69, *P* = 0.50; MF 0.80 vs pre-PMF 0.74, *P* = 0.96; PV 0.69 vs pre-PMF 0.74, *P* = 0.66).

These results revealed that a significant proportion of analyzed tiles with CIF scores assigned to bin 2 were not, in fact, derived from sample areas readily discernible by hematopathologists as equating to moderate / severe fibrosis amounting to WHO grade MF-2. This reflects the presence of microfoci or ‘hotspots’ of advanced fibrosis amidst non-fibrotic or less fibrotic areas. When this occurs, such fibrotic hotspots are typically deemed insufficient to warrant advanced grading by hematopathologists, or are either too small or too subtle to identify. Indeed, review of the CIF maps confirmed the presence of such microfibrotic hotspots throughout many MPN samples, most notably pre-PMF and PV.

### Cohort indexing of automated MPN fibrosis supports disease classification and assessment of disease progression

To enhance the visualization of our automated analysis, we performed principal component analysis (PCA) to create an abstracted 2-dimensional space that incorporates the average tile CIF score, tile distribution, and heterogeneity of tile distribution for our sample cohort (Figure 4A). As expected, PCA demonstrated clear separation of MF from reactive / normal and ET samples, with cases of PV seen to overlap each region in PCA space. The distribution of the PV samples in PCA space did not appear to be strongly associated with the JAK2 V617F variant allele frequency (supplemental Figure 7). The relationship between driver mutation status and PCA distribution for the ET, MF and pre-MF samples is shown in supplemental Figure 8.

**Figure 4.**
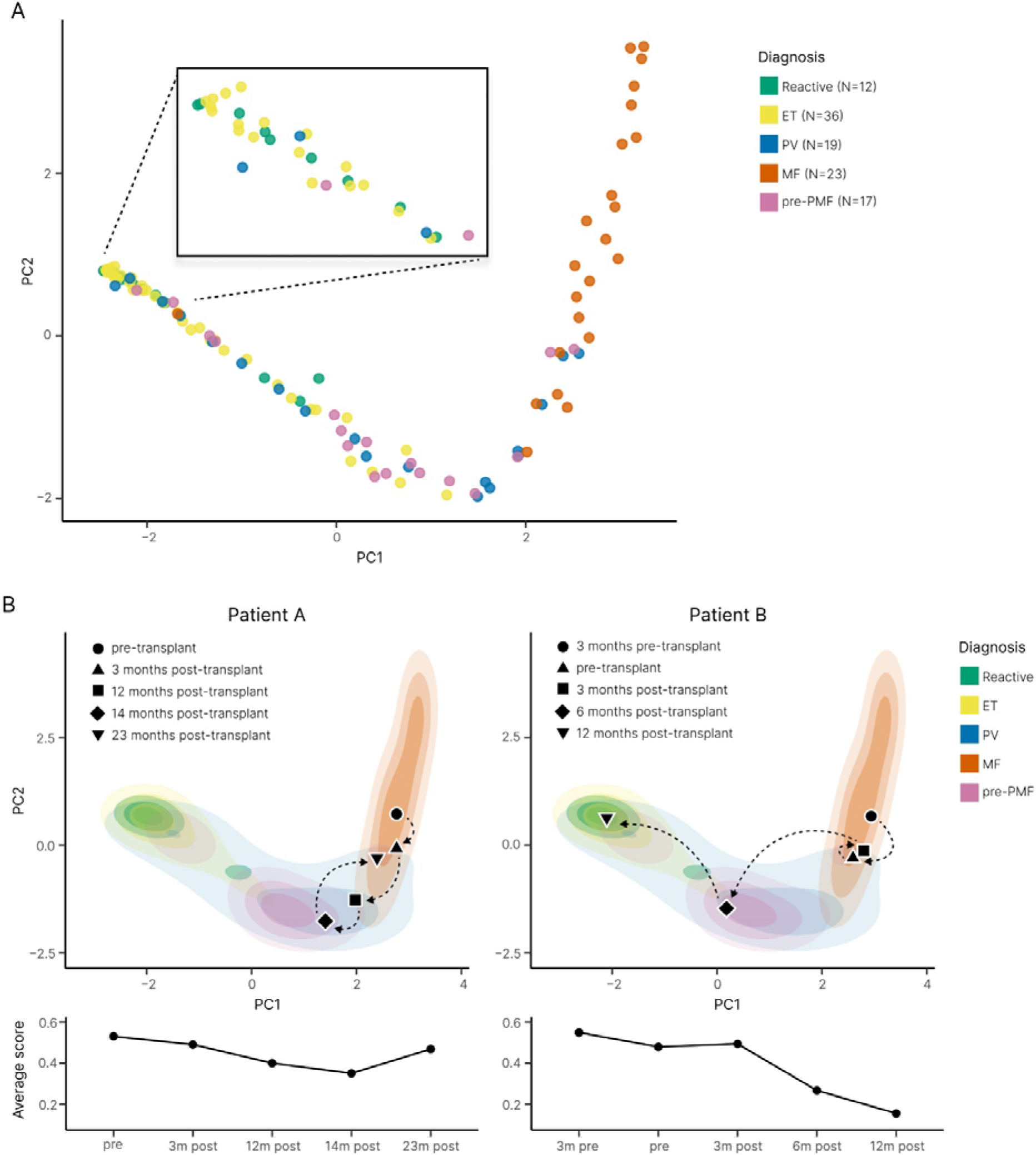
Disease cohort indexing discriminates between MPN samples and supports disease monitoring. (A) PCA plot of the abstract representations of sample fibrosis reveals the clustering of reactive cases and MPN disease subtypes. (B) Indexing the multivariable representation of fibrosis from individual patient samples to the PCA plot of the sample cohort enables monitoring of fibrotic progression. MF patients A and B underwent allogeneic bone marrow transplantation with subsequent marrow sampling to monitor disease response. Patient A demonstrated only modest improvement of marrow fibrosis post-transplant, with evidence of fibrotic relapse at 23 months (consistent with the results of clinical and laboratory disease monitoring). By contrast, Patient B demonstrated a profound reduction / normalization of marrow fibrosis within 12 months of transplantation, consistent with the clinical and laboratory findings.

**Figure 5.**
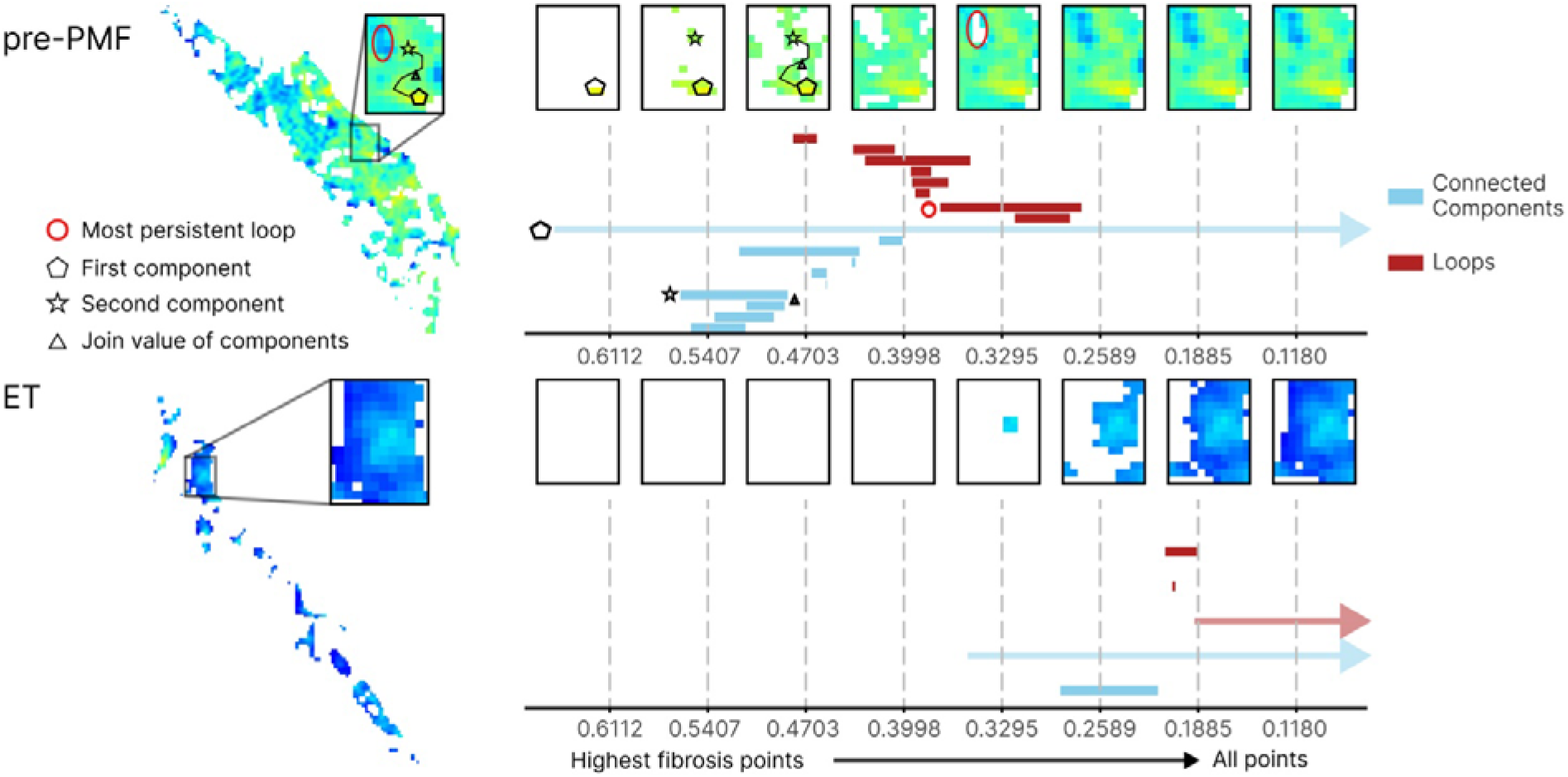
Topological data analysis (TDA) of fibrosis in ET and pre-PMF samples. Illustration of TDA of fibrotic foci in an ET and pre-PMF sample. Super-level set filtration on the images (left) allows computation of a topological fingerprint in the form of a barcode. The x-axis of the barcode corresponds to filtration values, i.e. fibrosis score thresholds, in the super-level set filtration. At high filtration values only the most fibrotic score pixels are included, with all points included at the lowest filtration values. The barcode in blue captures clusters or connected components of the hotspots (light blue bars) across the filtration; for high filtration values this corresponds to fibrosis hotspots which then merge with neighboring hotspots when all pixels between them are present in the filtration. Two examples of such hotspots in the filtration of a sample of pre-PMF are shown as a pentagon and a star, with the filtration value at which they merge highlighted by a triangle. In the barcode, we highlight the CIF values at which the hotspots are “born” in the corresponding bars with a pentagon and a star. When the pentagon and the star hotspots merge, the bar corresponding to the star hotspot ends (indicated by the triangle) and the now joint star-pentagon component continues to be monitored in the bar that corresponds to the pentagon hotspot. Infinite features, i.e. features that continue beyond the end of the filtration such as the clusters that consist of all fibrosis pixels in the sample once all pixels are included in the filtration, are represented as arrows extending beyond the x-axis. The barcode in red tracks one-dimensional holes (red bars) and their scale, also called persistence, in the images. The most persistent hole in the pre-PMF image is highlighted as a circle. We show the fibrosis score at which a hole appears (left end point) of the corresponding bar in the barcode with a circle.

Notably, when ET and pre-PMF samples were directly compared, both appeared to aggregate in distinct regions of the PCA plot with only minor overlap. Based on the PCA feature representations, we trained a random forest classifier to distinguish ET (n = 36) from pre-PMF (n = 17) samples; in three-fold cross validation (used to estimate the performance of a model by which data are split into 3 groups of approximately equal size) the classifier reached an area under the curve (AUC) of 0.71 for discriminating between these MPN subtypes (Figure 7B). Of note, two pre-PMF samples were seen to overlap with the PCA space primarily occupied by samples of MF, despite meeting WHO morphological diagnostic criteria including an overall WHO fibrosis grade of ≤ MF-1.

In addition to allowing a simplified assessment of the distribution of fibrosis within a cohort of reactive and MPN samples, PCA analysis allows changes in marrow fibrosis to be objectively detected and intuitively appreciated across sequential samples. This is of particular value in the interpretation of BMTs obtained from patients undergoing repeated biopsy to monitor disease response or progression (Figure 4B).

### Topological data analysis (TDA) of fibrotic features in ET and pre-PMF samples

Having identified significant differences in the average CIF tile score, tile distribution and heterogeneity between ET and pre-PMF, we sought to explore in more detail the fibrotic features associated with each subtype. We therefore extended our fibrotic feature analysis to include topological features as these have provided useful insight into other complex biomedical datasets^31-33,39^ (see Methods and supplemental Methods: Topological data analysis). The identified topological descriptors were combined with the original fibrotic features to train a random forest classifier (supplemental Figure 9; supplemental Table 9) with improved performance (AUC = 0.82 [combined TDA + original fibrotic features] vs AUC = 0.70 [original fibrotic features]). These topological differences corresponded to the structure of the fibrotic foci, with pre-PMF samples appearing to have a greater number of fibrotic foci. These foci also appeared to be more pronounced in relation to surrounding tissue areas when compared to ET. Notably, fibrotic foci in pre-PMF samples were also more likely to be connected by paths with high CIF scores when compared to ET, implying a potential spatial relationship between areas of advancing fibrosis in pre-PMF (Figure 5).

### Automated fibrosis analysis identifies patients at risk of fibrotic progression

Given the evidence of good disease separation of ET and pre-PMF samples in our local cohort using PCA, we hypothesized that our approach may allow improved early detection of MPN patients at risk of progression to secondary myelofibrosis. To evaluate this, we interrogated the PT-1 clinical trial cohort for patients diagnosed with ET in whom there was diagnostic evidence of transformation to secondary MF in the course of extended clinical follow-up. We identified 18 patients diagnosed with ET at trial enrollment in whom there was documented evidence of subsequent transformation to post-ET myelofibrosis (median days to transformation = 2356), and for whom we had access to analyzable pre-transformed reticulin-stained sections. As an internal control group we identified 17 PT-1 trial participants in whom there was no diagnostic evidence of transformation over a comparable or longer period of clinical follow-up (median follow-up = 4339 days). When indexed to the PCA plot derived from our locally sourced sample cohort (incorporating TDA), the PT-1 ET samples from non-transforming patients aggregated in the expected PCA space (Figure 6A). By contrast, around half (9 / 17) of the subsequently transformed PT-1 ET samples were seen to aggregate in the PCA space corresponding to cases of pre-PMF from our local cohort. A random forest classifier trained to discriminate between patients who did or did not subsequently transform to post-ET myelofibrosis achieved an AUC of 0.77 (Figure 6A; supplemental Table 10).

**Figure 6.**
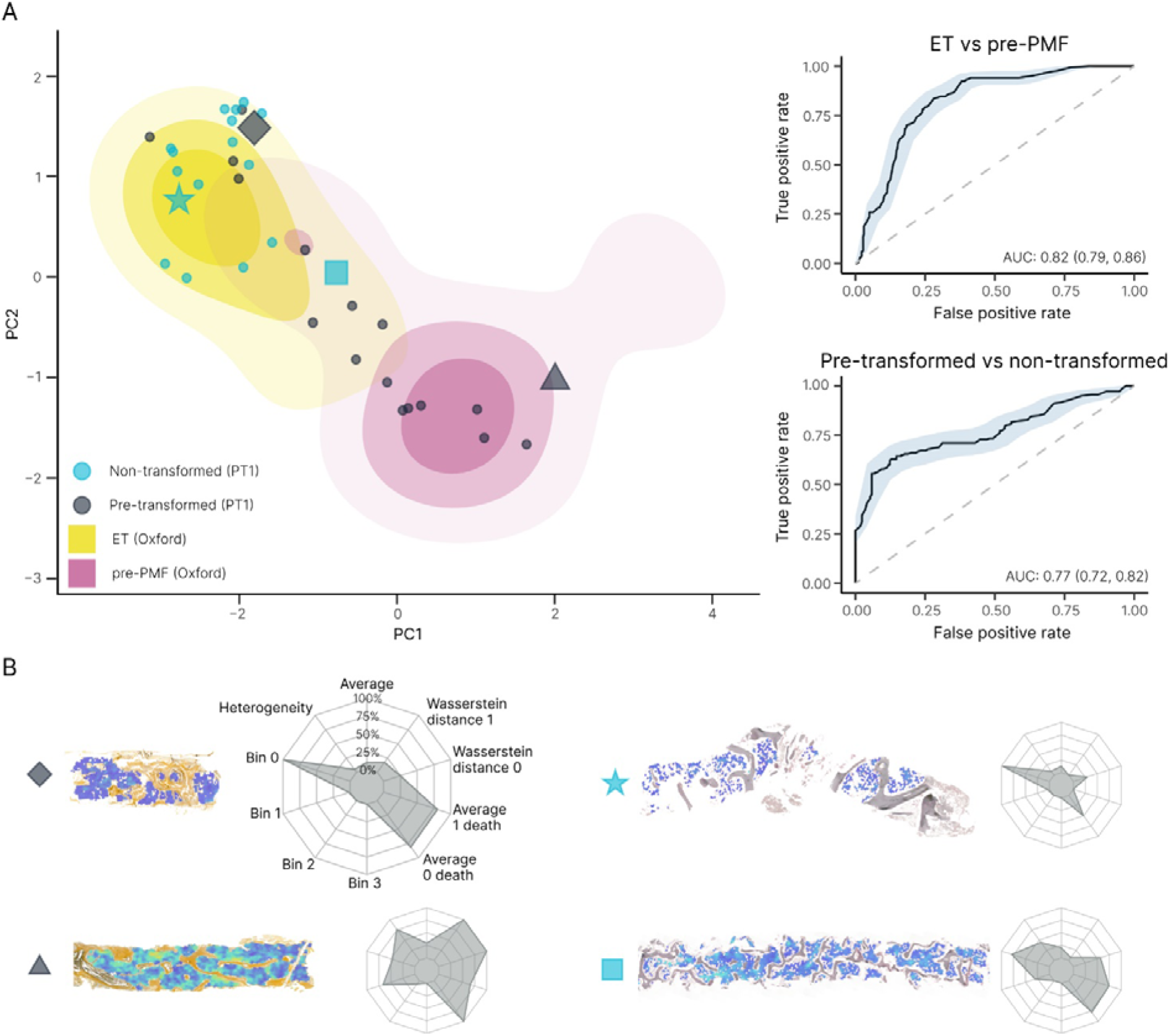
Combined fibrosis feature of ET, pre-PMF and PT-1 samples. (A) PCA plot of ET vs pre-PMF using fibrosis features (original + TDA) with overlay of pre-transformed and non-transformed cases of ET from the PT-1 cohort along with the ROC curves for ET vs pre-PMF and pre-transformed vs non-transformed PT-1 samples. (B) Examples of the CIF maps and corresponding radar plots of the analyzed samples.

**Figure 7.**
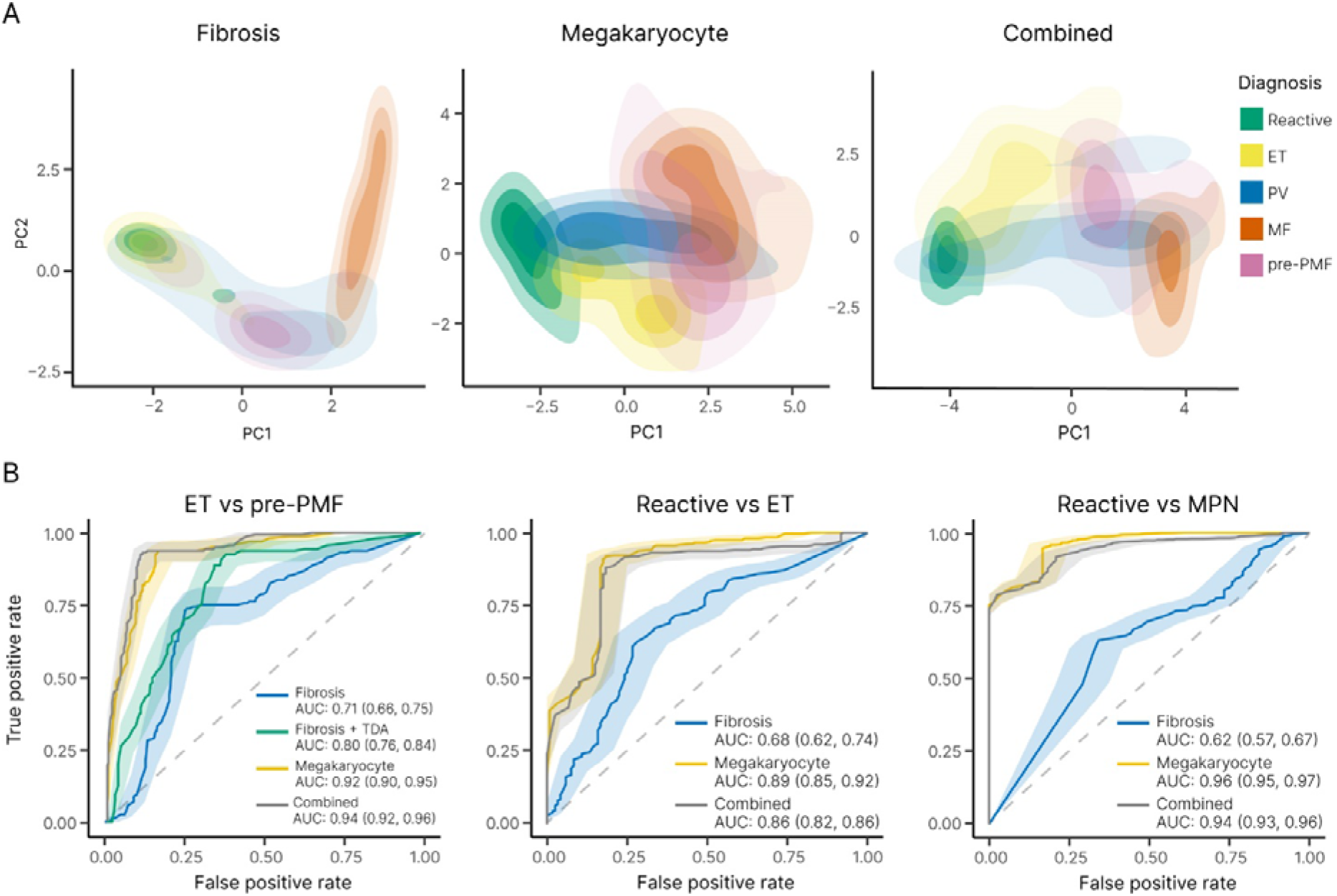
Combining fibrosis and megakaryocyte feature analysis improves the discrimination of MPN subtypes. (A) PCA plots of the fibrosis features, megakaryocyte features and combined features (fibrosis + megakaryocyte) reveals clustering of reactive samples and MPN subtypes. (B) Corresponding ROC curves (ET vs pre-PMF, reactive vs ET and reactive vs MPN) demonstrate the utility of combining fibrosis and megakaryocyte feature analysis in the assessment of MPNs, particularly for the discrimination of ET and pre-PMF samples (which includes TDA analysis of fibrotic features). For the purpose of comparison between fibrosis and megakaryocyte analyses for the ROC curve calculations, only samples for which both reticulin and H&E stains are available have been used (Reactive = 12, ET = 32, PV = 17, MF = 22, pre-PMF = 17).

### Integration of fibrotic and megakaryocytic features in MPN morphological assessment

Notwithstanding the diagnostic and prognostic potential of automated reticulin analysis, reticulin fibrosis is only one of several BMT features to be considered in the routine histological evaluation of MPNs. Indeed, in previous work employing ML to analyze megakaryocyte morphology and topology in BMTs we highlighted the potential of improved megakaryocyte analysis in the diagnosis and classification of MPNs^40^. Given the importance of both reticulin fibrosis and megakaryocyte analysis in MPN assessment, we sought to integrate both features using PCA in an attempt to improve the morphological resolution of MPN subtypes. When combined with our previous megakaryocyte feature PCA, fibrotic feature analysis demonstrated improved discrimination of ET and pre-PMF samples (AUC = 0.94 [megakaryocyte and fibrotic features] vs AUC = 0.92 [megakaryocyte features alone]) (Figure 7B; supplemental Table 9). By contrast, inclusion of fibrotic feature analysis did not enhance the discrimination of reactive and MPN (all subtypes) using megakaryocyte features (AUC = 0.94 [megakaryocyte and fibrotic features] vs AUC = 0.96 [megakaryocyte features alone]) (Figure 7B; supplemental Table 11), or the discrimination of reactive / normal and ET (AUC = 0.86 [megakaryocyte and fibrotic features] vs AUC = 0.89 [megakaryocyte features alone]) (Figure 7B; supplemental Table 12). This likely reflects the presence of variable amounts of minor fibrosis frequently encountered in healthy marrows, with significant feature overlap of non-fibrotic or mildly fibrotic MPN samples (Figure 4A; Figure 7A).

## Discussion

In this study we demonstrate that the complex and variable pattern of reticulin fiber deposition identified on routinely prepared reticulin-stained BMT slides can be captured on digital images and used to develop improved disease classification, grading and monitoring tools through image analysis technologies. We describe a set of computational methods designed to systematically capture the key morphological characteristics of fibrosis in MPNs and associate these features with particular MPN subtypes. This incorporates a platform that combines intuitive manual image handling tools with support from a ML model, thereby assisting hematopathologists in the efficient ranking of marrow fibrosis severity using conventional WHO criteria.

Objectively monitoring and quantitating fibrosis in BMTs is ideally suited for studies evaluating the effect of current therapies on disease progression in MPN, and for integration into future clinical trial designs evaluating novel therapeutic targets / drug candidates^41,42^. Without such approaches, incorporating marrow fibrosis assessment into robust clinical endpoints for the investigation of disease modifying agents in myelofibrosis will remain challenging.

The work presented here has significant potential in the challenging morphological assessment of patients in whom a differential diagnosis of ET and pre-PMF is considered. Using TDA we demonstrate that characteristic microfoci of advanced fibrosis are a recurrent feature of pre-PMF samples that are seldom encountered in ET. Detection and quantitation of these fibrotic microfoci is well beyond the scope of conventional histological assessment, and is not captured in current fibrosis grading classifications^11,18^. The clinical significance of this finding is supported by our retrospective analysis of samples obtained as part of the large PT-1 clinical trial of patients diagnosed with ET and receiving long term follow-up. TDA identified fibrotic features, similar to those observed in pre-PMF patients, in over half of those ET patients (for whom slides were available) who subsequently progressed to post-ET myelofibrosis while enrolled on the trial. Of note, patients eligible for PT-1 trial entry from 1997 to 2012 met the Polycythemia Study Group

Diagnostic criteria for ET, before widespread recognition of pre-PMF as a diagnostic category and formal adoption by the WHO in 2016^22-24^. This raises the possibility that at least a proportion of the ET patients subsequently transforming to secondary myelofibrosis might have had disease more in keeping with the current WHO entity of pre-PMF. Prospective studies determining the power of automated fibrosis assessment to predict myelofibrotic transformation in ET and pre-PMF classified using the latest WHO criteria are clearly indicated.

Our identification and description of fibrotic microfoci and related topological features within pre-PMF, and their association with fibrotic disease progression in ET, raises important questions about the factors driving early microfocal stromal fibrosis within the marrow. While the precise cellular mechanisms driving fibrosis in MPN remain to be elucidated, recent evidence from murine and human studies suggests that mal-differentiation of mesenchymal stem cells (MSC), driven by neoplastic HSCs and their inflammatory microenvironment, are a major determinant of distinct pre-fibrotic and fibrotic phases of disease^10,43^. The extent to which this process of stromal reprogramming is responsible for the microfoci of fibrosis identified in our current work clearly warrants further investigation. Moreover, the extent to which such early (potentially reversible) fibrotic foci may be important for widespread pathological changes in the surrounding marrow tissue, terminating in generalized marrow fibrosis, is also unclear. Intriguingly, analysis of the topological features embedded within our fibrosis data revealed not only increased numbers of fibrotic microfoci in pre-PMF samples when compared to ET, but also suggests that in pre-PMF these fibrotic hotspots are spatially related, possibly reflecting local conditioning of the surrounding stromal tissue that predispose to further foci of early fibrosis development. While speculative given the limited number of samples analyzed and the absence of prospective clinical data, this model of microfocal fibrotic progression in MPNs is entirely consistent with the growing body of evidence pointing to early HSC-driven abnormalities of the stem cell niche driving highly localized changes in the tissue microenvironment^8,9,44^.

Statistical descriptions of bone marrow morphological features using enhanced image analysis techniques have only recently been described, and application to fibrosis complements our recent work describing megakaryocyte morphology and topology in MPNs^40,45,46^. Of note, while the specific ML strategies employed for detecting and quantitating fibrosis in the form of a continuous score (CIF score) are distinct from those previously employed in our megakaryocyte analysis, they draw upon shared technical and infrastructural processes and deliver outputs that are readily integrated into shared analytical workflows. Indeed, we demonstrate how combining the morphological and topological features of fibrosis and megakaryocytes employed in conventional MPN diagnosis can be used to explore and refine our current understanding of disease boundaries. Importantly, we recognize that additional cellular and stromal morphological features are important in the current histological diagnosis and classification of MPNs, particularly cellular changes in non-megakaryocytic lineages and abnormalities of collagen deposition and bone. However, such features are in-turn well suited to novel ML approaches.

Fibrosis has long been recognised as an important pathological feature in diverse diseases affecting several organ systems such as liver, kidney and lung^47^. While improved measurements of histological fibrosis using image analysis / ML approaches have been particularly well described in liver disease^48,49^, our strategy of refining the topological features of fibrosis in the context of large curated patient cohorts and combining them with additional histological features is novel and has significant potential for rapid translation into other organ systems.

## Data Availability

All data produced in the present study are available upon reasonable request to the authors

## Data sharing statement

The datasets generated during and / or analyzed during the study are available from the corresponding author on request.

## Acknowledgements

The research was funded by a Cancer Research United Kingdom (CRUK) Early Cancer Detection Award and the National Institute for Health Research (NIHR) Oxford Biomedical Research Center (BRC). JR is supported through the EPSRC funded Seebibyte programme (EP/M013774/1). HAH gratefully acknowledges funding from EPSRC EP/K041096/1, EP/R005125/1 and EP/T001968/1, the Royal Society RGF\EA\201074 and UF150238, Leverhulme Trust and Emerson Collective. GG, BJS, HMB, and HAH are members of the Centre for Topological Data Analysis, funded by the EPSRC grant (EP/R018472/1). Computation used the Oxford Biomedical Research Computing (BMRC) facility, a joint development between the Wellcome Center for Human Genetics and the Big Data Institute supported by Health Data Research UK and the NIHR Oxford Biomedical Research Center. The PT-1 study was funded by the Medical Research Council, United Kingdom, and CRUK. The views expressed are those of the authors and not necessarily those of the NHS, the NIHR or the Department of Health. For the purpose of Open Access, the authors have applied a CC BY public copyright license to any Author Accepted Manuscript (AAM) version arising from this submission. We wish to express our gratitude to members of the 2019 CRUK Sandpit Team (HaemAI) for their support: Fayyaz Minhas, Wei Pang, Mathew Grech-Sollars, Peter R. Dunstan, and Alistair Easton.

## Authorship contributions

DR conceived and supervised the study with input from JR, BP, ALG and AJM. HR, KS, AA, DR and JR designed the study. HR, AA and KS designed the human-in-the-loop protocol for manual tile ranking; HR collected and summarized expert segmentations and manual ranking outputs. HR and KS implemented ML algorithms. DR, GDHT and GR verified ML predictions for fibrosis identification; HR and DR reviewed ML predictions for fibrosis grading. HR and KS performed statistical analyses. GG, HMB, HAH, and BJS designed the topological data analysis. GG performed topological computations and exploratory analysis. NS, BP, ALG, CNH and AJM identified samples and collected / provided access to clinical and genetic data. HR, KS and DR drafted the manuscript. All authors read and have given approval of the final manuscript.

