## Supplemental Material for "Continuous Indexing of Fibrosis (CIF): Improving the Assessment and Classification of MPN Patients"

### Methods

#### Fibrosis segmentation and manual rankings for training of the ranking-CNN model

We selected 18 BMT samples that spanned the range of WHO fibrosis grades MF-0 to MF-3 as per routine histological assessment. A hematopathologist (DR) segmented tissue areas corresponding to each MF grade. Invalid areas, such as bone and crush artifacts were also segmented to exclude them from analysis and subsequent training of the fibrosis ML model (supplemental Figure 3A). After manual segmentation, we extracted uniformly sized image tiles (512 x 512 pixels [0.22 𝜇m per pixel]) labeled with the corresponding MF grade (supplemental Figure 3B). We then applied feature similarity to identify tile pairs with high similarity^1^ (supplemental Figure 3C), and asked three hematopathologists (DR, GDHT, GR) to label which of the two tiles was more fibrotic (supplemental Figure 3D). We used these labeled pairs to train the initial ranking model (supplemental Figure 3E). We repeated this process iteratively with a human-in-the-loop approach, where subsequent training rounds of the ranking model use tile pairs derived from the previous round, with manual pairwise ranking performed by three hematopathologists.

For each round of model training, we divide the ranked list of tiles into four bins based upon their predicted score (bin 0 = lowest scores; bin 3 = highest scores). We further divide each bin into two subgroups, one comprising the highest scores within the bin and the other comprising the lowest scores. To create tile pairs for each bin we selected one tile each from the highest and lowest scoring subgroups. We ensured tile pairs with subtle differences in fibrosis severity were included. The number of pairs for the training and validation set are shown in supplemental Table 4.

The ranking-CNN model architecture consists of six convolutional layers and one fully connected layer followed by batch normalization, rectified linear unit (ReLU), and max pooling with the filter size of two. The first convolutional layer has a filter size of five. Other convolutional layers have a filter size of three and a stride of one. The output layer outputs a score for a given input. The structure of the ranking-CNN model follows a Siamese network which consists of two identical neural networks with shared parameters and weights^2^. During the backpropagation, the networks use the same initialization and gradient. The model is implemented using Python and the deep learning Pytorch library^3^. The details of the model hyperparameters are shown in supplemental Table 3.

For each pair of inputs x_A_ and x_B_, the model generates corresponding output scores y_A_ and y_B_. The input which produces the higher output score is of higher rank (i.e. has more severe fibrosis). During training, the model tries to minimize the loss where the given input pair produces output scores that are in the incorrect order when compared to ground truth. To do this, the Sigmoid function (eq. 1) converts the difference O_AB_ = y_A_-y_B_ to the probability ($P_{AB}$) that x_A_ is more severe than x_B_, for example, if $P_{AB}=0.7$, indicates there is a 70% chance that x_A_ is more severe than x_B_. The ground truth values would be $\bar{P}_{AB}=1.0$ if the left tile is more severe and $\bar{P}_{AB}=0.0$ if the right tile is more severe. The probability distribution is then used to calculate the loss using the Cross-Entropy (CE) which measures how close two probability distributions (true probability distribution and predicted probability distribution) are (eq. 2).

$$Sigmoid:$$

$P_{AB}= \frac{e^{o_{AB}}}{1+e^{o_{AB}}}$	**(1)**

$$Cross Entropy Loss:$$

| $L_{AB}=-\bar{P}_{AB}logP_{AB}-(1-\bar{P}_{AB})log(1-P_{AB})$  $=-\bar{P}_{AB}\{logP_{AB}+log(1-P_{AB})\}-log(1-P_{AB}$)  $= -\bar{P}_{AB}o_{AB}+log(1+e^{O_{AB}})$ | $\bar{P}_{AB}$ : True probability  $P_{AB}$: Predicted Probability | **(2)** |
| --- | --- | --- |

Once the loss is calculated we update the network weights via backpropagation. We use model prediction accuracy (percentage of candidate pairs correctly ranked) as the evaluation metric. Additionally, in order to ensure the model is generalisable we increase the variability in the training data through several augmentation techniques; horizontal and vertical flips, rotation (-45⁰ to +45⁰), and shear (-10 to +10).

#### Bone segmentation model training

Intact and fragmented trabecular or cortical bone is a prominent feature of many BMT samples. In order to identify bone in new samples we trained a separate segmentation model. First, a hematopathologist (DR) manually segmented bone structures from 89 samples (62 for training; 27 for validation). Then we extracted tiles 512 x 512 pixels [1.76 𝜇m per pixel] at 5X magnification. The bone segmentation model architecture is based on UNet^4^ which comprises an encoder network (similar to the CNN model in Ranking-CNN) followed by a decoder network which restores the condensed feature map into the original size of the input image tile. The output is a segmentation map defined by a prediction for each pixel. The hyperparameter settings are provided in supplemental Table 2.

#### Topological data analysis

To quantify the spatial distribution of foci in the predicted fibrosis heatmap, we compute topological features such as connected components (i.e. foci and more general fibrotic regions), which we refer to as *0-dimensional* features, and holes (i.e. voids or loops inside the components), which we refer to as *1-dimensional* features, as the image is thresholded by a range of fibrosis scores. For each image, we take a super-level set filtration of the tiles by CIF score (see Figure 5). A super-level set filtration consists of a stack of binary images, thresholded (from below): the first step of the filtration includes the tiles with the highest fibrosis score pixel, the next filtration step continues to include the next highest fibrosis score pixel, and continues until all fibrosis score pixels are included. Next, we compute topological features (the *homology*) of this filtration of spatial fibrosis scores. Specifically, we track the evolution of fibrotic components and holes appearing and merging / disappearing into other features through the filtration (i.e. as the threshold decreases to include more pixels of the data). Each feature is summarized by a threshold interval on which the feature is visible, and the set of all intervals, called the *persistence barcode*^5^ or fibrotic fingerprint (Figure 5), provides a signature of the image. Therefore the computational output of each sample is a multiset of intervals, where the left endpoint represents the fibrosis score at which a fibrotic feature appears (“birth” threshold) and the right endpoint is the fibrosis score value at which it disappears or ‘dies’ (“death” threshold); long bars or intervals represent a large range of fibrotic scores for which the feature persists.

While many vectorisation methods can be applied to barcodes to make them suitable for machine learning^6,7^, we chose interpretable barcode statistics that were particularly well-suited to pre-PMF / ET classification. We extracted the statistics from exploratory analysis using a random forest classifier consisting of 100 decision trees, from the Python package scikit-learn^8^. Starting with a feature vector including average fibrosis, bin values, heterogeneity (Shannon entropy), and 30 topological features, we evaluated the topological statistics using Gini importance^9^, improvement to the cross-validation score of our random forest classifier between the ET and pre-PMF class (supplemental Figure 9), and correlation with the non-spatial features (supplemental Figure 10). The initial topological features under consideration included the following, computed using Python Ripser version 0.6.2^10^:

- birth times and persistence of the top 5 classes of connected components (by birth); (10)
- birth times and persistence of the top 5 classes of loops (by birth); (10)
- the Wasserstein 2-norm of the persistence barcodes for connected components and loops; (2)
- the persistent entropy of the persistence barcodes for connected components and loops; (2)
- number of features of the persistence barcodes for connected components and loops; (2)
- number of features and mean center of mass of the persistence barcodes; (4)

Upon analysis, the features with highest Gini importance, highest marginal contribution to cross-validation score, and lowest correlation with non-spatial features were: the average death threshold for 0- and 1-dimensional features (normalized by average fibrosis level of the image); and the Wasserstein norm of the 0- and 1-dimensional barcodes (normalized by image size). The normalized death times can be interpreted as the relative height of saddle points and local minima in the image, and correspond to properties of the links between foci. The Wasserstein norms can be viewed as a measure of overall topological heterogeneity.

##### Classification

We trained a random forest classifier with 100 estimators (the nodes are expanded until all leaves contain less than two samples) using Gini impurity^11^ to split each node to discriminate between ET (n = 36) and pre-PMF (n = 17) samples, reactive (n = 12) and MPN (n = 98) samples, and between reactive (n = 12) and ET (n = 36) samples. We used three-fold cross-validation; in each cross-validation the ratio of each subtypes are preserved to maintain the ratio at their original levels. As the performance metrics, classification accuracy (each class and overall), precision, recall, F1-score, and area under the receiver operating characteristic curve (AUC) were recorded for each cross-validation and overall performance. We used the Python package scikit-learn^8^ to apply a random forest classifier and calculate the performance metrics.

### Supplemental Tables

**Supplemental Table 1. Clinicopathological summary of the local (Oxford) sample cohort.**

|  |  | **Overall** | **Reactive** | **ET** | **PV** | **MF** | **pre-PMF** |
| --- | --- | --- | --- | --- | --- | --- | --- |
|  |  | n = 107 | n = 12 | n=36 | n=19 | n=23 | n=17 |
| **Blood counts** | |  |  |  |  |  |  |
| Platelet count |  | n=79 | NA | n=35 | n=10 | n=17 | n=17 |
| (10^9^ / L) | Median (range) | 523  (121, 1397) |  | 602  (121, 1285) | 474  (234, 697) | 290  (132, 832) | 826  (622, 1397) |
| White cell count |  | n=79 | NA | n=35 | n=10 | n=17 | n=17 |
| (cells / L) | Median (range) | 8.3  (4.7, 59) |  | 7.89  (4.7, 30.9) | 8.65  (5.3, 13.5) | 13.8  (4.7, 59) | 9.1  (4.75, 33.55) |
| Hemoglobin |  | n=79 | NA | n=35 | n=10 | n=17 | n=17 |
| (g / L) | Median (range) | 13.4  (7.5, 117) |  | 13.8  (10, 17) | 15.3  (11.2, 19.6) | 9.8  (7.5, 17.3) | 13.5  (8.4, 117) |
| **Mutation status** | |  | | | | | |
| TN | | 8 | NA | 8 | 0 | 0 | 0 |
| JAK2 (V617F) | | 64 | NA | 23 | 18 | 14 | 9 |
| CALR | | 15 | NA | 3 | 0 | 5 | 7 |
| MPL | | 4 | NA | 1 | 0 | 2 | 1 |

**Supplemental Table 2. Hyperparameters of the bone segmentation model.**

| **Parameter** | **Value** |
| --- | --- |
| Batch Size | 50 |
| Learning Rate | 0.0001 |
| Optimizer | ADAM |
| Loss | Pixel-wise Cross Entropy Loss |

**Supplemental Table 3. Hyperparameters of the ranking-CNN model.**

| **Parameter** | **Value** |
| --- | --- |
| Batch Size | 30 |
| Learning Rate | 0.0001 |
| Optimizer | ADAM |
| Loss | Cross Entropy Loss |

**Supplemental Table 4. Training and validation samples & image tiles for the ranking-CNN model.**

|  | Training (# of tiles) | Validation (# of tiles) |
| --- | --- | --- |
| Initial round | 11 (3503) | 6 (1880) |
| 1st round | 14 (2479) | 6 (596) |
| 2nd round | 14 (2325) | 6 (587) |
| **Total** | 39 (8307) | 18 (3063) |

**Supplemental Table 5**. **Manual pairwise ranking concordance of three pathologists across image tile pair sets (%)**.

| **MF Grade Comparison** | **Concordance (%)** |
| --- | --- |
| 0 vs 0 | 100 |
| 0 vs 1 | 97.80 |
| 0 vs 2 | 99.56 |
| 0 vs 3 | 100 |
| 1 vs 1 | 69.08 |
| 1 vs 2 | 82.24 |
| 1 vs 3 | 91.74 |
| 2 vs 2 | 86.92 |
| 2 vs 3 | 70.05 |
| 3 vs 3 | 72.22 |
| **Total** | **88.40** |

**Supplemental Table 6. Ranking performance within different image pairs & interobserver agreement (%) based on the graded tiles.**

|  | **GT vs Prediction** | **H1 vs GT** | **H2 vs GT** | **H3 vs GT** | **H1 vs Prediction** | **H2 vs Prediction** | **H3 vs Prediction** | **H1 vs H2** | **H1 vs H3** | **H2 vs H3** |
| --- | --- | --- | --- | --- | --- | --- | --- | --- | --- | --- |
| **0vs0** | 100 | 100 | 100 | 100 | 100 | 100 | 100 | 100 | 100 | 100 |
| **0vs1** | 98.53 | 98.9 | 99.63 | 99.27 | 98.9 | 98.9 | 98.53 | 98.53 | 98.17 | 98.9 |
| **0vs2** | 100 | 100 | 100 | 99.56 | 100 | 100 | 99.56 | 100 | 99.56 | 99.56 |
| **0vs3** | 100 | 100 | 100 | 100 | 100 | 100 | 100 | 100 | 100 | 100 |
| **1vs1** | 88.82 | 86.84 | 92.11 | 90.14 | 76.97 | 86.18 | 88.16 | 78.95 | 76.97 | 82.24 |
| **1vs2** | 89.96 | 94.98 | 91.89 | 95.37 | 88.8 | 88.03 | 90.73 | 86.87 | 90.35 | 87.26 |
| **1vs3** | 95.87 | 98.76 | 97.11 | 95.87 | 95.45 | 93.08 | 95.04 | 95.87 | 94.63 | 92.98 |
| **2vs2** | 95.33 | 97.2 | 95.33 | 94.39 | 92.52 | 94.39 | 93.46 | 92.52 | 91.59 | 89.72 |
| **2vs3** | 79.19 | 90.36 | 91.88 | 87.82 | 72.59 | 75.13 | 80.2 | 82.23 | 78.17 | 79.7 |
| **3vs3** | 90 | 88.89 | 91.11 | 92.22 | 85.56 | 87.78 | 82.22 | 80.00 | 81.11 | 83.33 |

*H1 / H2 / H3 = individual hematopathologists; GT = ground truth (initial manual segmentation)*

**Supplemental Table 7. Average ranking agreement of model prediction vs hematopathologists (H) & interobserver agreement based on the graded tiles.**

|  | **H vs Prediction** | **Interobserver** |
| --- | --- | --- |
| **0vs0** | 100 | 100 |
| **0vs1** | 98.78 | 98.53 |
| **0vs2** | 99.85 | 99.71 |
| **0vs3** | 100 | 100 |
| **1vs1** | 83.77 | 79.39 |
| **1vs2** | 89.19 | 88.16 |
| **1vs3** | 94.52 | 94.49 |
| **2vs2** | 93.46 | 91.28 |
| **2vs3** | 75.97 | 80.03 |
| **3vs3** | 85.19 | 81.48 |
| **Average** | 92.07 | 91.31 |

**Supplemental Table 8. Average fibrosis feature values for MPN and reactive samples.**

|  | **Average CIF Score** | **Heterogeneity** | **Bin 0 (%)** | **Bin 1 (%)** | **Bin 2 (%)** | **Bin 3 (%)** |
| --- | --- | --- | --- | --- | --- | --- |
| **Reactive** | 0.17 | 0.26 | 90.58 | 7.95 | 1.46 | 0.00 |
| **ET** | 0.19 | 0.36 | 82.36 | 16.10 | 1.51 | 0.03 |
| **PV** | 0.30 | 0.69 | 46.93 | 35.20 | 16.99 | 0.88 |
| **MF** | 0.55 | 0.80 | 6.17 | 15.80 | 52.54 | 25.48 |
| **pre-PMF** | 0.30 | 0.74 | 46.52 | 38.85 | 14.16 | 0.47 |

**Supplemental Table 9. Classification performance of random forest classifier for ET and pre-PMF samples.**

| **Classification Accuracy** | | | | |
| --- | --- | --- | --- | --- |
|  | **Fold 1** | **Fold 2** | **Fold 3** | **Overall** |
| **Fibrosis only** | | | | |
| **ET** | 0.80 | 0.82 | 0.73 | 0.78 |
| **pre-PMF** | 0.74 | 0.60 | 0.48 | 0.60 |
| **Overall** | 0.78 | 0.75 | 0.64 | 0.72 |
| **Fibrosis + TDA** | | | | |
| **ET** | 0.80 | 0.72 | 0.89 | 0.81 |
| **pre-PMF** | 0.70 | 0.60 | 0.55 | 0.61 |
| **Overall** | 0.77 | 0.69 | 0.77 | 0.74 |
| **Megakaryocyte only** | | | | |
| **ET** | 0.90 | 0.82 | 0.96 | 0.89 |
| **pre-PMF** | 0.80 | 0.6 | 0.95 | 0.79 |
| **Overall** | 0.87 | 0.75 | 0.96 | 0.86 |
| **Fibrosis + TDA + Megakaryocyte** | | | | |
| **ET** | 0.92 | 0.82 | 1.00 | 0.91 |
| **pre-PMF** | 0.86 | 0.60 | 0.93 | 0.81 |
| **Overall** | 0.90 | 0.75 | 0.98 | 0.88 |
| **AUC, Precision, Recall and F1** | | | | |
|  | **Fold 1** | **Fold 2** | **Fold 3** | **Overall** |
| **Fibrosis only** | | | | |
| **AUC (95% CI)** | 0.77 (0.70, 0.85) | 0.71 (0.61, 0.80) | 0.66 (0.58, 0.74) | 0.71 (0.66, 0.75) |
| **Precision** | 0.75 | 0.71 | 0.61 | 0.69 |
| **Recall** | 0.77 | 0.71 | 0.61 | 0.69 |
| **F1** | 0.76 | 0.71 | 0.61 | 0.69 |
| **Fibrosis + TDA** | | | | |
| **AUC (95% CI)** | 0.87 (0.82, 0.92) | 0.71 (0.62, 0.79) | 0.85 (0.79, 0.91) | 0.80 (0.76, 0.84) |
| **Precision** | 0.74 | 0.65 | 0.76 | 0.71 |
| **Recall** | 0.75 | 0.66 | 0.72 | 0.71 |
| **F1** | 0.74 | 0.65 | 0.73 | 0.71 |
| **Megakaryocyte only** | | | | |
| **AUC (95% CI)** | 0.96 (0.94, 0.99) | 0.87 (0.81,0.93) | 1.0 (0.99, 1.0) | 0.92 (0.90, 0.95) |
| **Precision** | 0.85 | 0.71 | 0.95 | 0.84 |
| **Recall** | 0.85 | 0.71 | 0.96 | 0.84 |
| **F1** | 0.85 | 0.71 | 0.96 | 0.84 |
| **Fibrosis + TDA + Megakaryocyte** | | | | |
| **AUC (95% CI)** | 0.98 (0.97, 1.00) | 0.86 (0.81, 0.92) | 1.00 (1.00, 1.00) | 0.94 (0.92, 0.96) |
| **Precision** | 0.89 | 0.71 | 0.98 | 0.86 |
| **Recall** | 0.89 | 0.71 | 0.97 | 0.86 |
| **F1** | 0.89 | 0.71 | 0.97 | 0.86 |

**Supplemental Table 10. Classification performance of random forest classifier for pre-transformed and non-transformed PT-1 trial ET samples.**

| **Classification Accuracy** | | | | |
| --- | --- | --- | --- | --- |
|  | **Fold 1** | **Fold 2** | **Fold 3** | **Overall** |
| **Fibrosis only** | | | | |
| **Non-trans** | 0.70 | 0.70 | 0.50 | 0.63 |
| **Pre-trans** | 0.67 | 0.53 | 0.67 | 0.62 |
| **Overall** | 0.68 | 0.62 | 0.58 | 0.63 |
| **Fibrosis + TDA** | | | | |
| **Non-trans** | 0.98 | 0.83 | 0.68 | 0.82 |
| **Pre-trans** | 0.50 | 0.80 | 0.67 | 0.66 |
| **Overall** | 0.72 | 0.82 | 0.68 | 0.74 |
| **AUC, Precision, Recall and F1** | | | | |
|  | **Fold 1** | **Fold 2** | **Fold 3** | **Overall** |
| **Fibrosis only** | | | | |
| **AUC (95% CI)** | 0.87 (0.80, 0.93) | 0.74 (0.65, 0.83) | 0.82 (0.74, 0.89) | 0.78 (0.74, 0.83) |
| **Precision** | 0.68 | 0.62 | 0.59 | 0.63 |
| **Recall** | 0.68 | 0.62 | 0.58 | 0.63 |
| **F1** | 0.68 | 0.61 | 0.58 | 0.63 |
| **Fibrosis + TDA** | | | | |
| **AUC (95% CI)** | 0.80 (0.71, 0.88) | 0.86 (0.80, 0.93) | 0.67(0.57, 0.78) | 0.77 (0.72, 0.82) |
| **Precision** | 0.79 | 0.82 | 0.68 | 0.75 |
| **Recall** | 0.74 | 0.82 | 0.68 | 0.74 |
| **F1** | 0.71 | 0.82 | 0.67 | 0.74 |

**Supplemental Table 11. Classification performance of random forest classifier for reactive samples and MPN subtypes.**

| **Classification Accuracy** | | | | |
| --- | --- | --- | --- | --- |
|  | **Fold 1** | **Fold 2** | **Fold 3** | **Overall** |
| **Fibrosis only** | | | | |
| **Reactive** | 0.25 | 0.10 | 0.00 | 0.12 |
| **MPN** | 0.93 | 0.93 | 1.00 | 0.96 |
| **Overall** | 0.85 | 0.84 | 0.88 | 0.86 |
| **Megakaryocyte only** | | | | |
| **Reactive** | 0.70 | 0.48 | 1.00 | 0.73 |
| **MPN** | 1.00 | 1.00 | 0.93 | 0.97 |
| **Overall** | 0.96 | 0.94 | 0.93 | 0.95 |
| **Fibrosis + Megakaryocyte** | | | | |
| **Reactive** | 0.68 | 0.48 | 0.10 | 0.42 |
| **MPN** | 1.00 | 1.00 | 0.94 | 0.98 |
| **Overall** | 0.96 | 0.94 | 0.84 | 0.91 |
| **AUC, Precision, Recall and F1** | | | | |
|  | **Fold 1** | **Fold 2** | **Fold 3** | **Overall** |
| **Fibrosis only** | | | | |
| **AUC (95% CI)** | 0.82 (0.77, 0.87) | 0.61 (0.53, 0.70) | 0.44 (0.37, 0.51) | 0.62 (0.57, 0.67) |
| **Precision** | 0.62 | 0.53 | 0.44 | 0.57 |
| **Recall** | 0.59 | 0.52 | 0.50 | 0.54 |
| **F1** | 0.6 | 0.52 | 0.47 | 0.54 |
| **Megakaryocyte only** | | | | |
| **AUC (95% CI)** | 1.0 (1.0, 1.0) | 0.90 (0.86,0.94) | 0.99 (0.99, 1.0) | 0.96 (0.95, 0.97) |
| **Precision** | 0.98 | 0.97 | 0.82 | 0.88 |
| **Recall** | 0.85 | 0.74 | 0.96 | 0.85 |
| **F1** | 0.90 | 0.81 | 0.87 | 0.86 |
| **Fibrosis + Megakaryocyte** | | | | |
| **AUC (95% CI)** | 1.00 (0.99, 1.00) | 0.91 (0.87, 0.94) | 0.89 (0.86, 0.92) | 0.94 (0.93, 0.96) |
| **Precision** | 0.98 | 0.97 | 0.53 | 0.83 |
| **Recall** | 0.84 | 0.74 | 0.52 | 0.7 |
| **F1** | 0.89 | 0.81 | 0.52 | 0.74 |

**Supplemental Table 12. Classification performance of random forest classifier for reactive and ET samples.**

| **Classification Accuracy** | | | | |
| --- | --- | --- | --- | --- |
|  | **Fold 1** | **Fold 2** | **Fold 3** | **Overall** |
| **Fibrosis only** | | | | |
| **Reactive** | 0.25 | 0.08 | 0.00 | 0.11 |
| **ET** | 0.68 | 0.79 | 0.83 | 0.77 |
| **Overall** | 0.56 | 0.60 | 0.61 | 0.59 |
| **Megakaryocyte only** | | | | |
| **Reactive** | 0.98 | 0.50 | 1.00 | 0.83 |
| **ET** | 1.00 | 0.93 | 0.8 | 0.9 |
| **Overall** | 0.99 | 0.81 | 0.85 | 0.88 |
| **Fibrosis + Megakaryocyte** | | | | |
| **Reactive** | 0.93 | 0.50 | 0.73 | 0.72 |
| **ET** | 1.00 | 0.95 | 0.81 | 0.92 |
| **Overall** | 0.98 | 0.83 | 0.79 | 0.86 |
| **AUC, Precision, Recall and F1** | | | | |
|  | **Fold 1** | **Fold 2** | **Fold 3** | **Overall** |
| **Fibrosis only** | | | | |
| **AUC (95% CI)** | 0.56 (0.46, 0.65) | 0.81 (0.72, 0.91) | 0.77 (0.71, 0.83) | 0.68 (0.62, 0.74) |
| **Precision** | 0.47 | 0.41 | 0.35 | 0.42 |
| **Recall** | 0.47 | 0.43 | 0.41 | 0.44 |
| **F1** | 0.47 | 0.42 | 0.38 | 0.43 |
| **Megakaryocyte only** | | | | |
| **AUC (95% CI)** | 1.00 (1.00, 1.00) | 0.79 (0.71, 0.87) | 0.97 (0.95, 0.99) | 0.89(0.85, 0.92) |
| **Precision** | 1.00 | 0.78 | 0.82 | 0.85 |
| **Recall** | 0.99 | 0.71 | 0.9 | 0.86 |
| **F1** | 0.99 | 0.73 | 0.83 | 0.85 |
| **Fibrosis + Megakaryocyte** | | | | |
| **AUC (95% CI)** | 1.00 (1.00, 1.00) | 0.78 (0.70, 0.86) | 0.82 (0.75, 0.89) | 0.86 (0.82, 0.86) |
| **Precision** | 0.98 | 0.82 | 0.74 | 0.83 |
| **Recall** | 0.96 | 0.73 | 0.77 | 0.82 |
| **F1** | 0.97 | 0.75 | 0.75 | 0.82 |

### Supplemental Figures

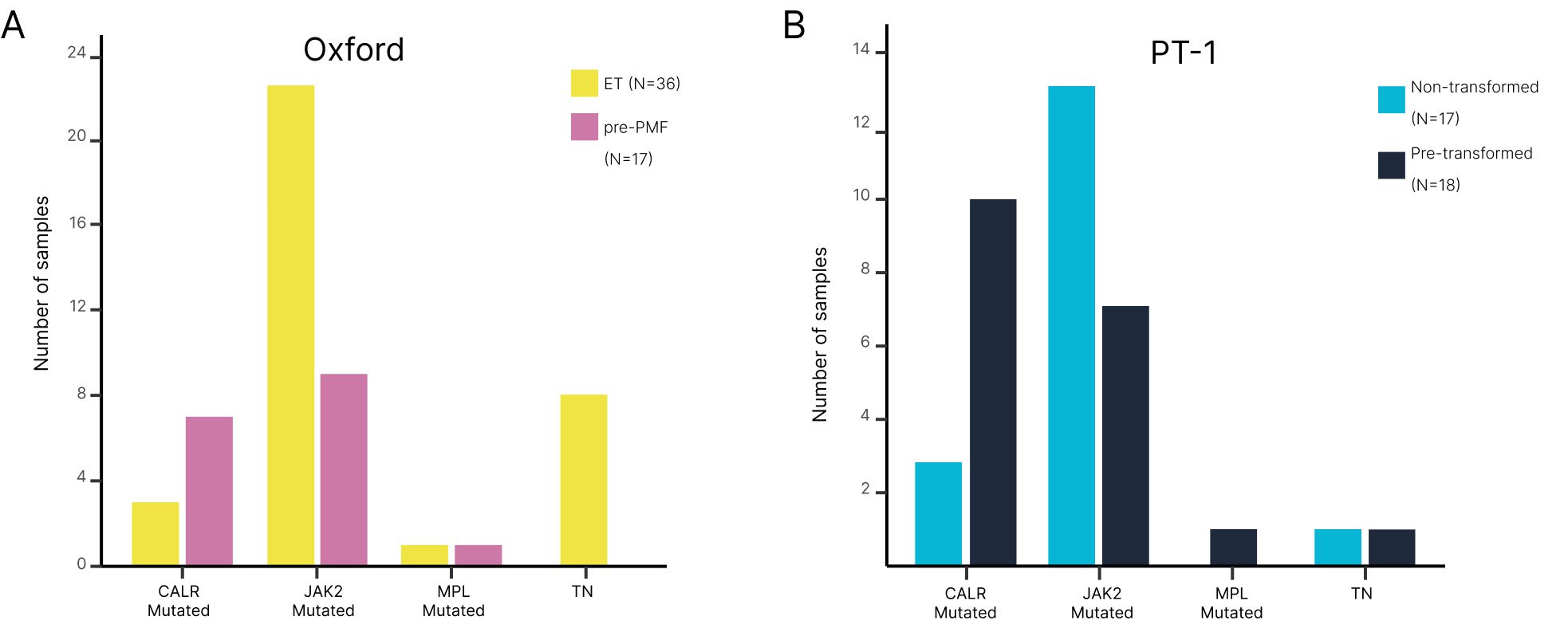

**Supplemental Figure 1**. **Distribution of driver mutation status within (A) ET and pre-PMF samples from the Oxford cohort, and (B) pre-transformed and non-transformed ET samples from the PT-1 cohort.**

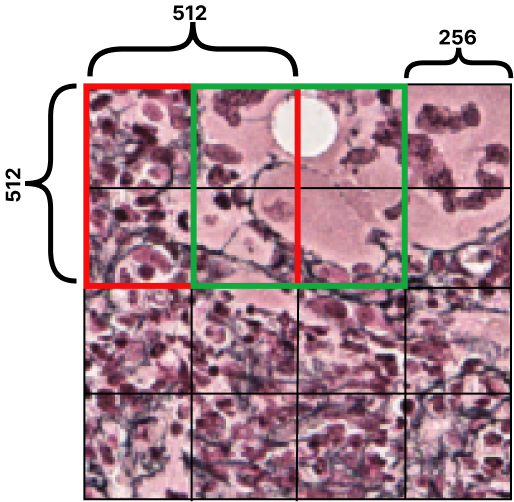

**Supplemental Figure 2.** **Example of the sliding window approach with a stride of 256 pixels (0.22 𝜇m per pixel) to extract the image tiles (512 x 512 pixels [0.22 𝜇m per pixel]).** In this example, this image field will contribute a total of 9 tiles for analysis.

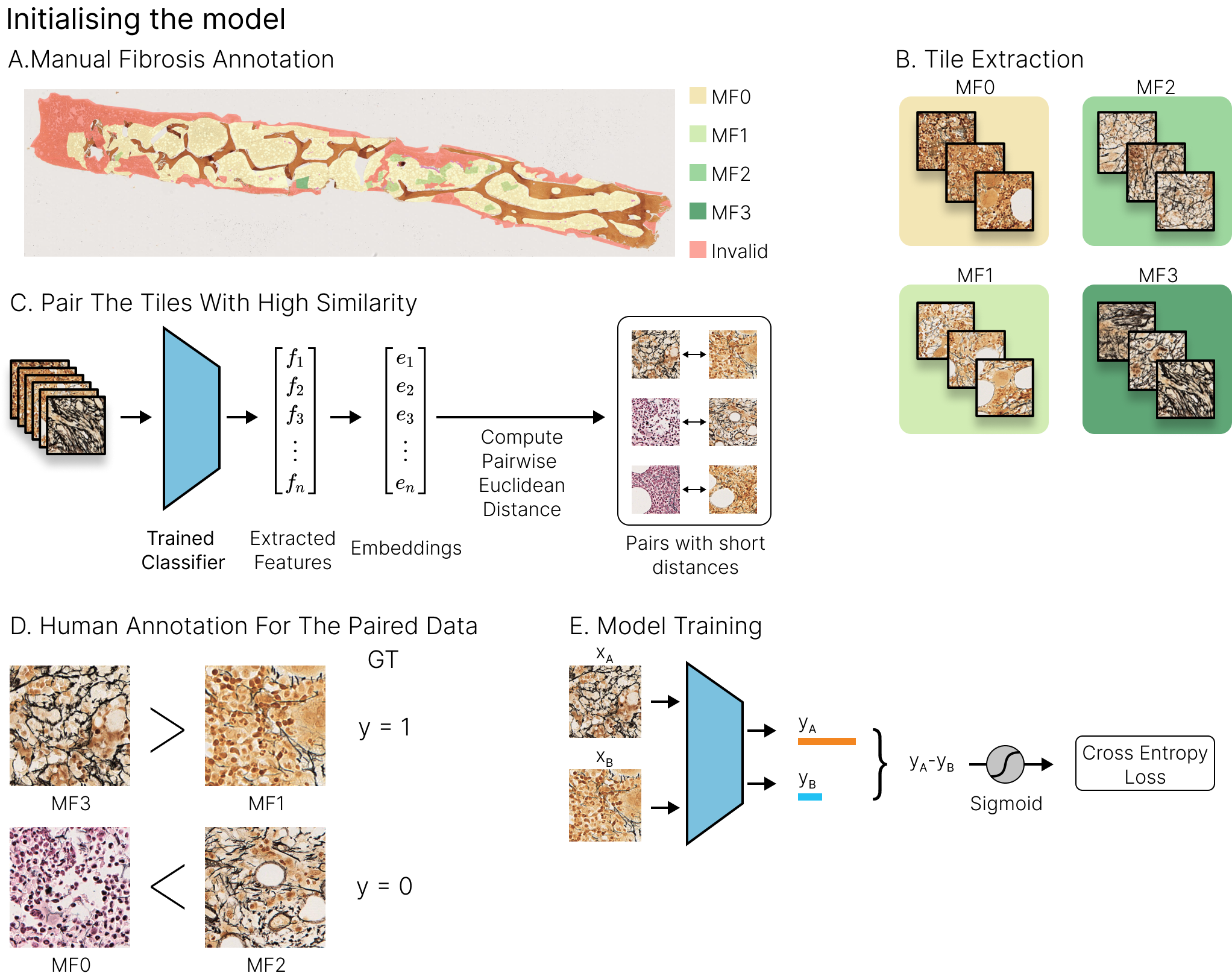

**Supplemental Figure 3**. **Overview of the initial tile pair acquisition and model training**. (A) An unlabeled sample is manually segmented for areas of each fibrosis (MF) grade. (B) Tiles are extracted from the segmented regions and labeled with the corresponding MF grade. (C) Tiles with high similarity are paired using Euclidean distance. (D) Manual ranking of tile pairs. (E) The Ranking-CNN model is trained on the set of ranked pairs.

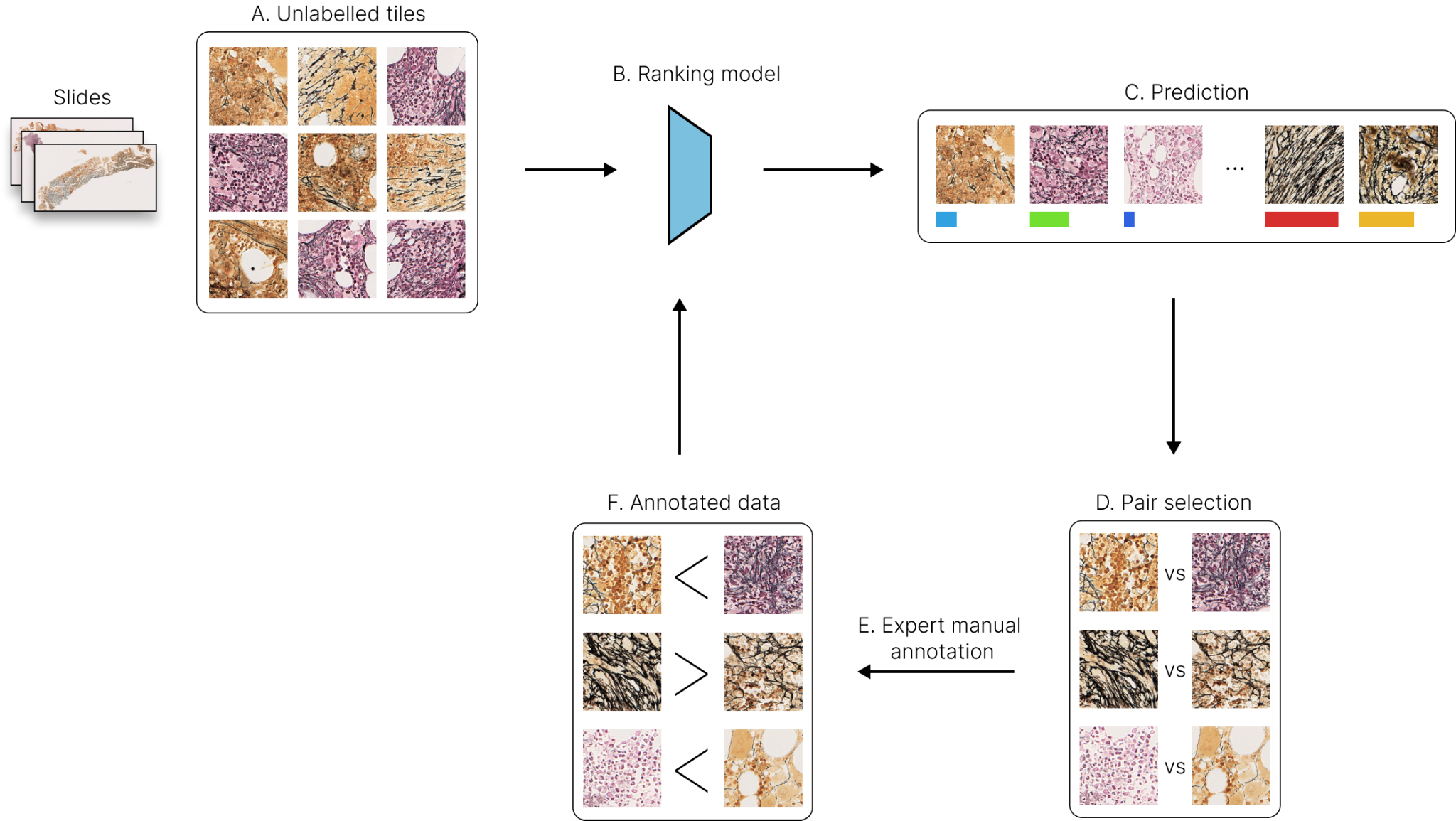

**Supplemental Figure 4. Overview of the human-in-the-loop approach for manual ranking**. (A) From the unlabeled sample, tiles are extracted from the regions of interest. (B, C) The trained ranking-CNN model predicts the score of the unlabeled tiles. (D) Based on the predicted rank, similar pairs are selected for manual ranking. (E, F) Manually ranked pairs are used for subsequent rounds of model training.

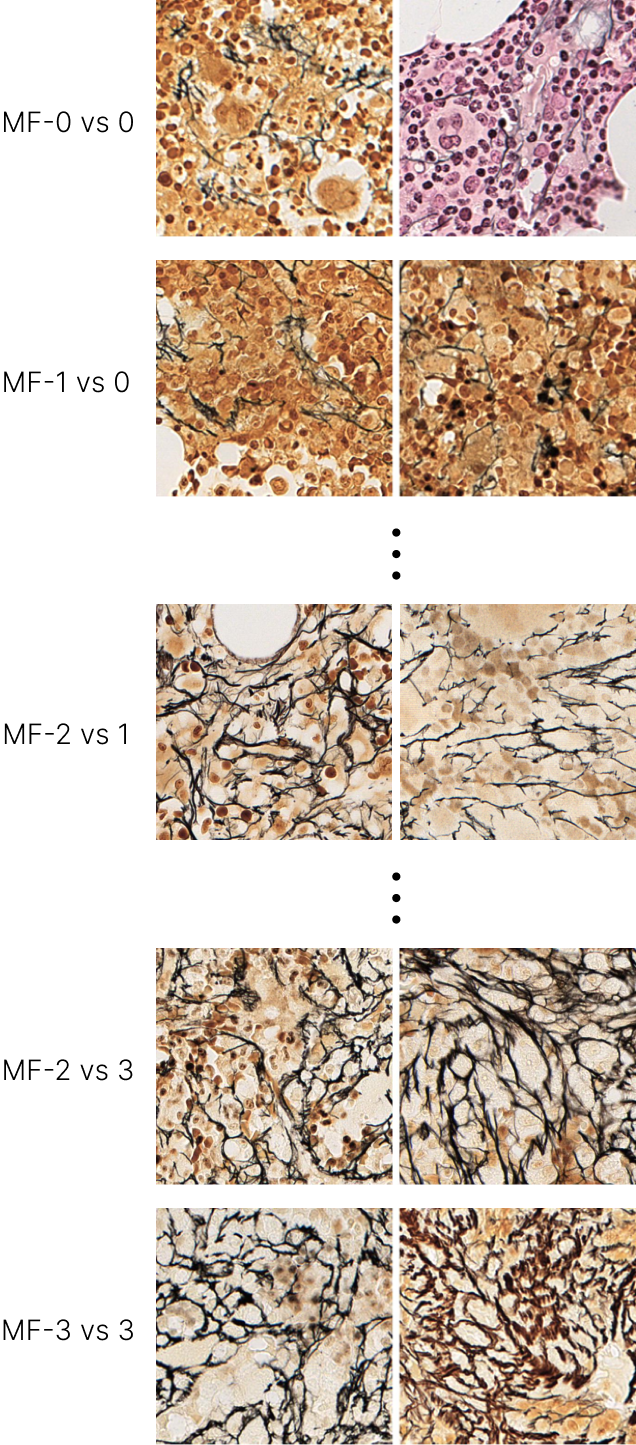

**Supplemental Figure 5. Examples of image tile pairs from each fibrosis (MF) grade.**

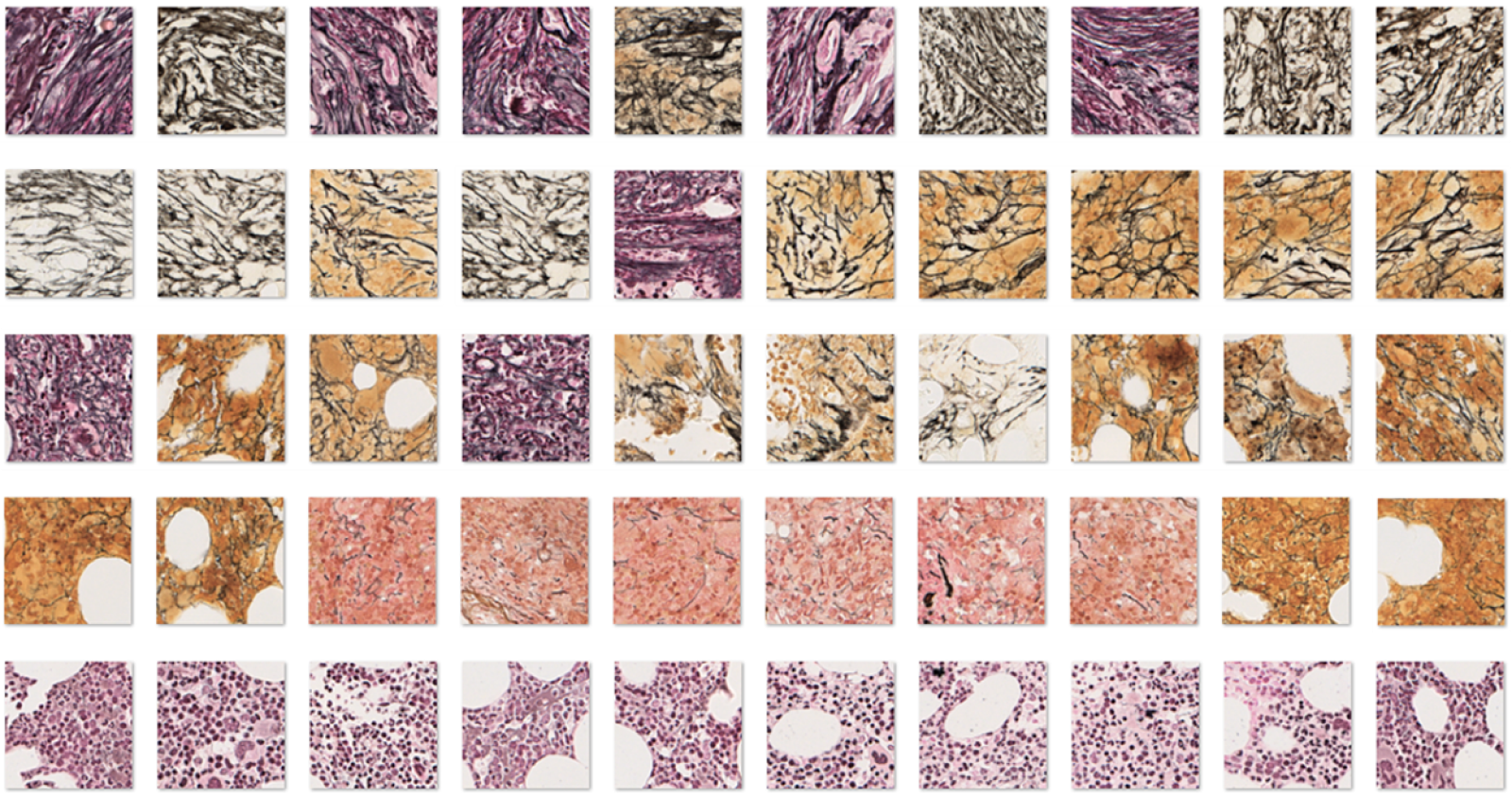

**Supplemental Figure 6. Examples of the predicted ranking of a subset of image tiles from the validation set**. Images from the top row have the most extensive fibrosis, with thick reticulin fibers and numerous intersections. The bottom row of images have virtually no detectable fibrosis, with intervening rows having progressively less fibrosis from top to bottom.

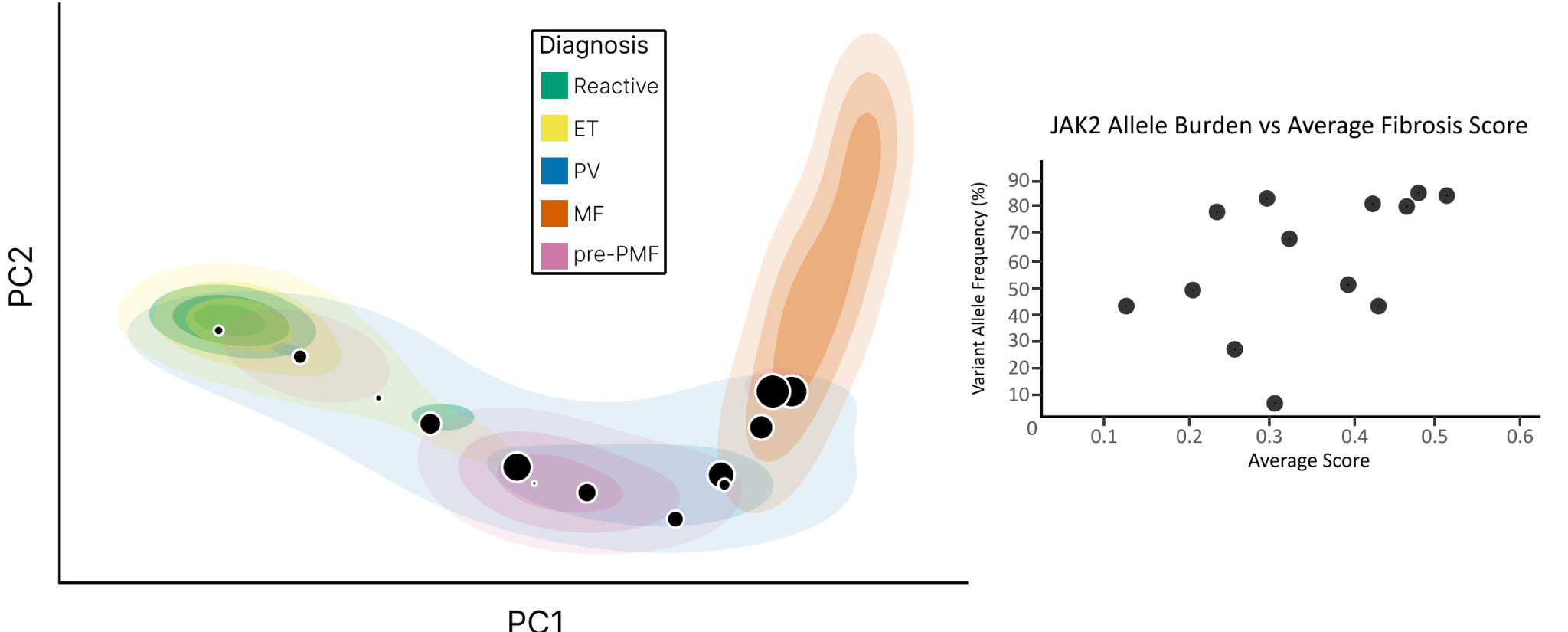

**Supplemental Figure 7. Association between JAK2 V617F variant allele frequency (VAF) and location on fibrosis PCA space for samples of PV.** Circles corresponding to each PV sample are shown, with circle size proportionate to the JAK2 VAF on the PCA plot.

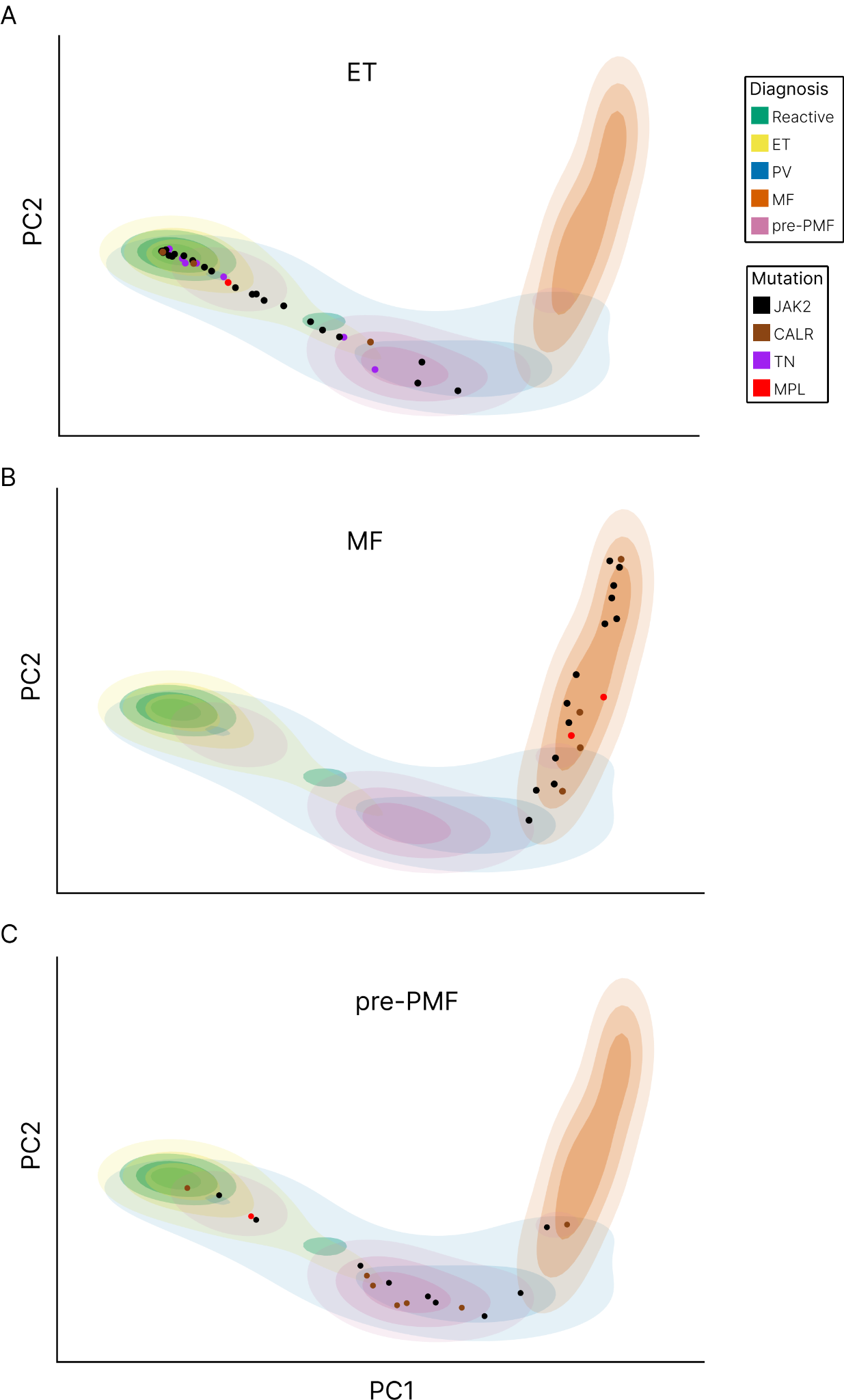

**Supplemental Figure 8.** **Association between driver mutation status and location on fibrosis PCA space for samples of ET, MF and pre-PMF (A-C).**

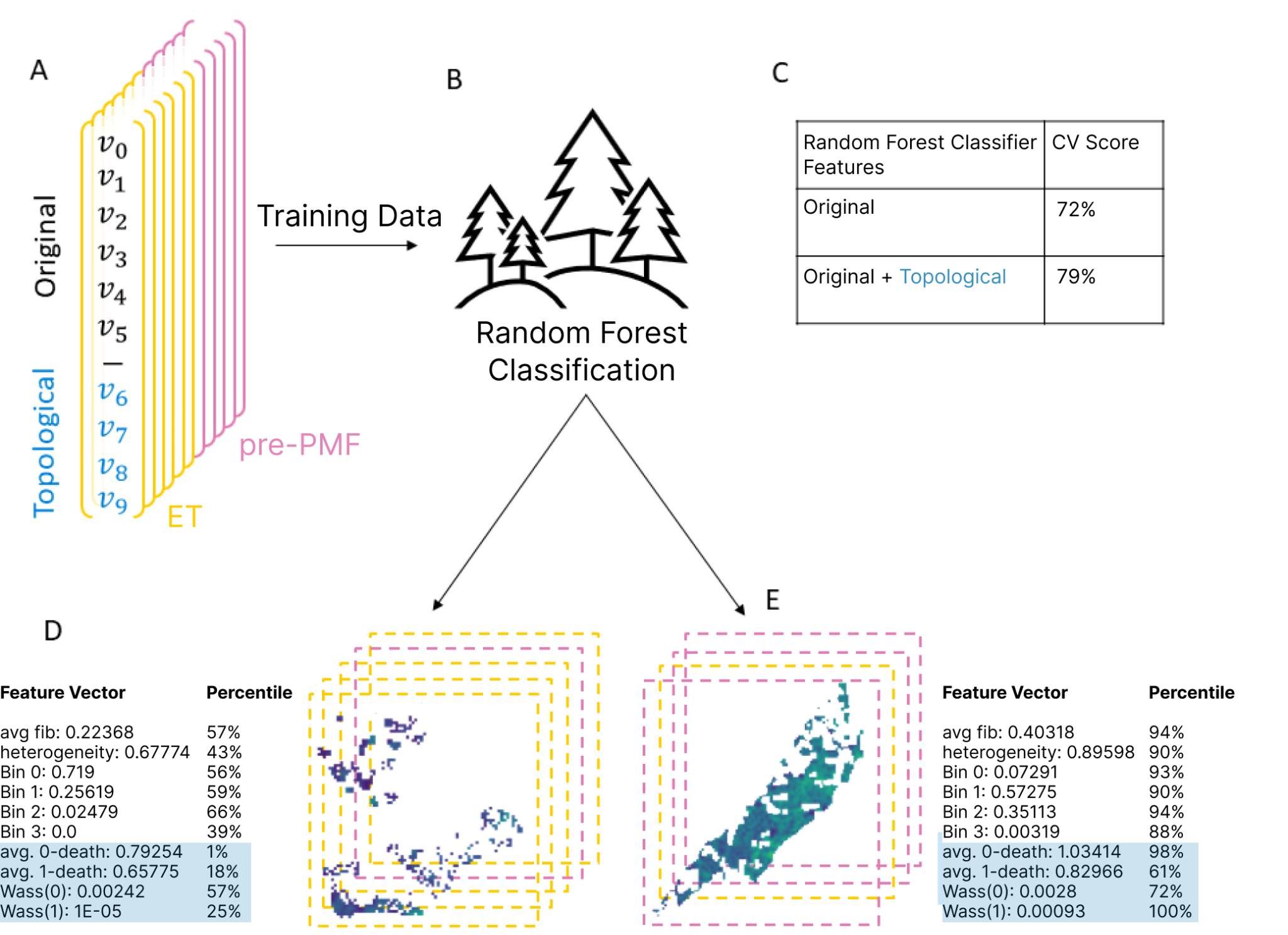

**Supplemental Figure 9.** **Overview of the fibrosis feature selection using the random forest classifier**. (A) Fibrosis-related features (Original: average CIF score, heterogeneity, Bin 0, Bin 1, Bin 2, Bin 3 and Topological [TDA]: avg 0-death, avg 1-death, Wass(0) and Wass(1) extracted from ET and pre-PMF samples. (B) Training of random forest classifier based on the extracted features. (C) Classifier accuracy of original features vs original + topological (TDA) features. (D) Example of the extracted features from ET and (E) pre-PMF samples with the corresponding CIF score maps.

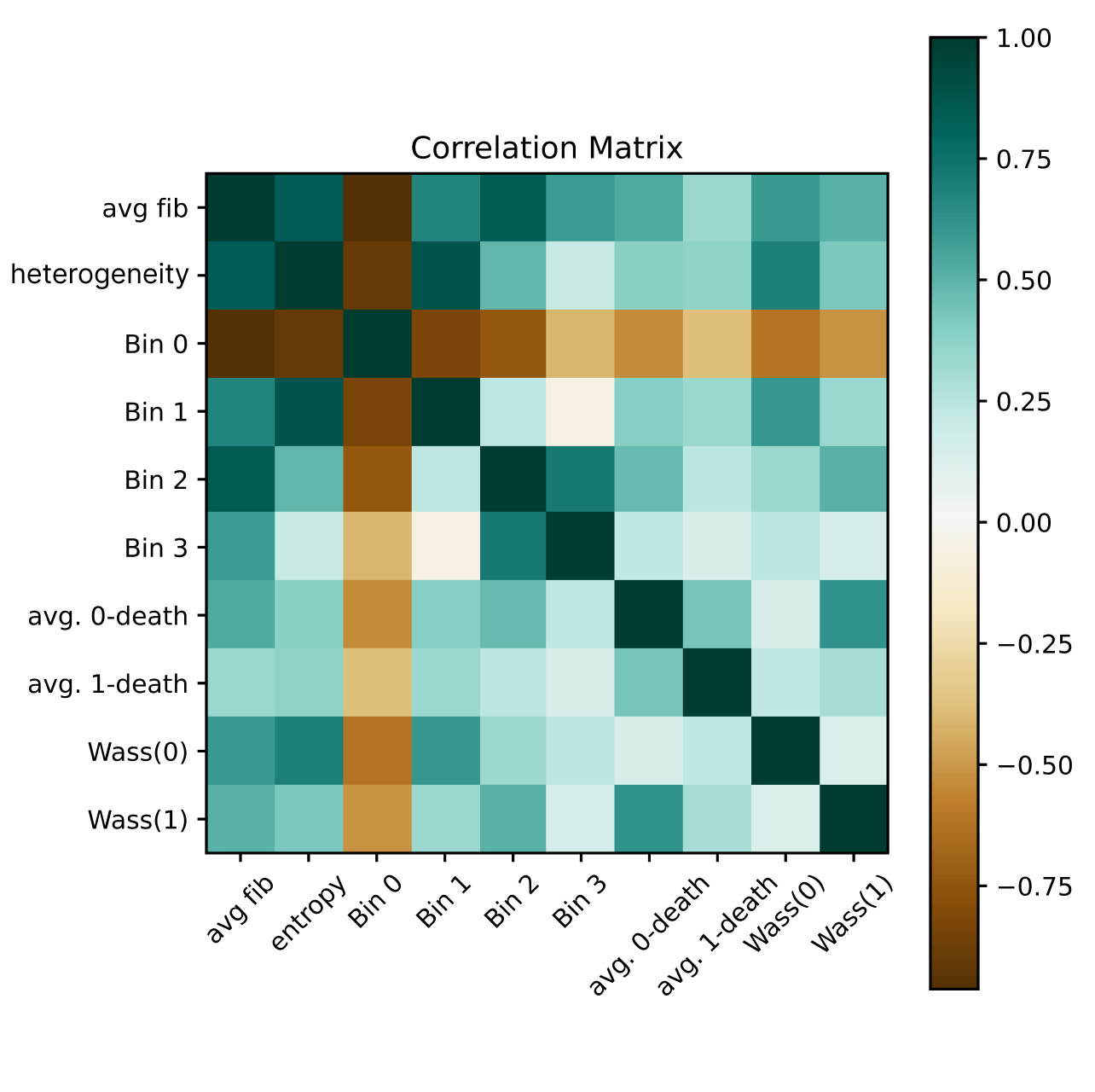

**Supplemental Figure 10**. **Correlation matrix based on the extracted features (original + topological [TDA]).** Pearson product moment correlation was calculated between vectors consisting of extracted features across all images.
